# Pilot long-read amplicon sequencing of clinical malaria infections from the R21/Matrix-M Phase III vaccine trial

**DOI:** 10.64898/2026.11.24.26368765

**Authors:** Lydia J. Jennings, Annie J. Forster, James Docker, Katerina Rapi, Amy Flaxman, Gabrielle Rockett, Yanxia Wu, Amanda Chong, Sophie Weston, Benedict Hollingdale, Patrick Trimby-Smith, Amelia Bajer, Jason Hendry, Emma Beaumont, Thomas Roberts, Paolo Piazza, Alassane Dicko, Mainga Hamaluba, Ally Olotu, Jean-Bosco Ouédraogo, Hamtandi M. Natama, Halidou Tinto, Lisa Stockdale, Adrian V. S. Hill, Alexander J. Mentzer, Mehreen S. Datoo, Gavin Band

## Abstract

With the rollout of effective liver-stage vaccines against *Plasmodium falciparum* (*P. falciparum*) malaria across sub-Saharan Africa, a major concern is that parasite populations might evolve genetic forms of vaccine resistance. Here, we describe a pilot project to sequence the *PfCSP* gene, which encodes the major immunological target of the currently approved vaccines, and nine other gene regions in infections of children in the R21/Matrix-M vaccine Phase III trial. Our approach uses DNA extracted from study dried blood spots, and adapts the recently-developed NOMADS long-read amplicon panel to the PacBio platform. We develop a pipeline to call full-length amino acid sequences and infer polyclonality, and apply this to a single clinical infection from each of 319 children recruited at age 5-17 months, from four study sites in Burkina Faso, Mali, and Kenya. Approximately half of these infections are inferred to be polyclonal; we observed lower levels of polyclonality in infections of R21-vaccinated individuals, and in individuals who experienced fewer episodes in the year following vaccination. At *CSP*, we find that no study haplotype perfectly matches the full *CSP* reference coding sequence, and only 8% fully match the vaccine-encoded C-terminal sequence. The majority of haplotype variation was observed in the central NANP/NVDP repeat (37-46 repeats), in the Th2R and Th3R epitope regions, and at a common 19-residue deletion variant. No major differences in variant frequencies were observed between R21-vaccinated and control participants, but we caution that these pilot data are not sufficiently powered to detect plausible true vaccine escape effects. Our data provide a new resource of *P. falciparum* haplotype data across *CSP* and multiple other loci in these populations, and we illustrate this by analysing antimalarial resistance profiles in detail. The methods described here are now being applied across more than 4,500 trial timepoints in the R21/Matrix-M Phase III study.

## Introduction

Despite advances in vector control, diagnosis, and antimalarial therapies since the 1990s, malaria remains a major global health problem with the highest burden of disease and mortality falling on young children in Africa. The introduction of effective vaccines including RTS,S/AS01 (here referred to as RTS,S) and R21/Matrix-M (R21) – both of which have been recommended by the World Health Organisation (WHO) and are being implemented globally – has the potential to substantially reduce disease burden and should accelerate progress toward eradication. As deployment at population scale begins, however, there is great interest in understanding population variation in vaccine responses. A particularly important question is whether parasite genetic diversity may influence vaccine performance, or drive parasite evolutionary responses caused by vaccine-induced selection pressure.

Both RTS,S and R21 target the parasite circumsporozoite protein (CSP), a key surface antigen expressed during the pre-erythrocytic stage of the parasite lifecycle. CSP is expressed on the surface of sporozoites (the parasite form transmitted by mosquitos and which initially invade liver cells) and on the surface of infected hepatocytes(1). CSP can therefore induce both B- and T-cell mediated immune responses in the host, and phase III trials of both vaccines have shown efficacy in large population cohorts(2, 3).

The R21 and RTS,S vaccine sequence includes the full CSP C-terminal as well as a set of 18 repeats of an NANP motif, that encode key antibody epitopes. These were chosen in part because they appear conserved across malaria species(4). The underlying gene (*CSP*) is, however, highly genetically variable, with considerable variation observed in the number of repeat segments and in the C-terminal sequence. Prior evidence of the RTS,S/AS01 Phase III trial suggests that *CSP* genetic variation could modulate vaccine efficacy(5), suggesting higher efficacy against parasites carrying *CSP* alleles matching the vaccine sequence compared to heterologous variants. Reanalyses of the same Phase III dataset have reached similar conclusions(6–8). However, this finding has not been replicated in independent datasets, including in a RTS,S/AS01 Phase IIb trial(9) which was not well powered to detect this effect. It is therefore of great interest to extend these analyses to other large datasets where vaccine-induced immune response and efficacy can be assessed.

Here we work with infections ascertained during the large R21/Matrix-M Phase III trial(2), describing a pilot study to analyse parasite variation from dried blood spots (DBS) from four study sites. We develop methodology using long-read sequencing of gene amplicons to analyse *CSP* and nine other gene regions using the PacBio platform. For this pilot, we apply this to a single timepoint of clinical infection – the last such infection within 12 months of vaccination - from 319 children aged 5-17 months at time of recruitment. The resulting data constitute a new resource of haplotype variation in these populations from 2021-2022 and we use this to analyse patterns of polyclonality and genetic diversity in *CSP*, as well as examining the presence of antimalarial drug-resistance haplotypes across loci. A proof-of-principle test for evidence of association between CSP amino acid variation and vaccine group does not find any strong relationship, but we highlight that data from the full study will be needed to fully assess this.

## Results

### Overview and sample selection

This pilot project aimed to characterise *Plasmodium falciparum* parasite genetic variation in a subset of infections observed in the first 12 months of the R21/Matrix-M Phase III trial(2). **Figure 1** outlines the overall workflow and sample numbers used in our study. In brief, a total of 4,644 children aged 5-36 months were included in the trial modified per-protocol analysis, of whom a third were vaccinated with R21 and the remainder with a control rabies vaccine (Abhayrab) in a randomised framework(2). Among these children, approximately a quarter (N=1,157) experienced clinical infection events in the first 12 months post-vaccination and were treated according to local guidelines. For this pilot we focussed on the subset of children in the younger age category (5-17 months old at time of vaccination) and on the last clinical infection timepoint experienced by each child within this 12-month period, if any. Infections from a total of N=425 children met these inclusion criteria (Methods) and of these, DBS from 372 samples were physically identified within the collection and taken forward for analysis. To help assess replicability we also included a second sample (ggN-2f5-Re9) for one individual, taken one day after the initial infection and showing low parasitaemia (referred to as a non-clinical repeat sample below; **Supplementary Table 1**).

**Figure 1.**
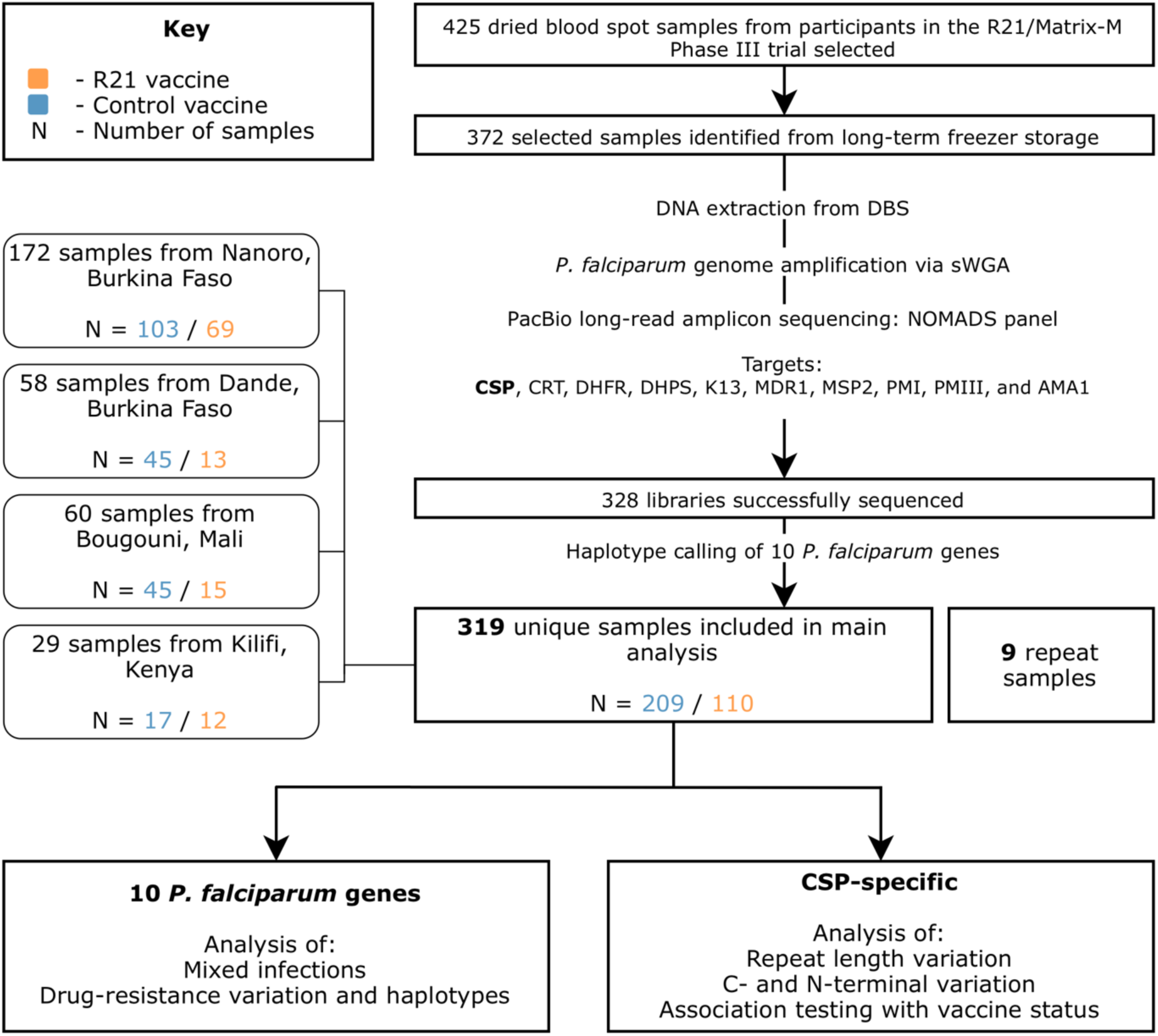
Flowchart showing the workflow from sample selection to final analysis and sample counts. Orange and blue numbers indicate infections of R21- and control-vaccinated participants respectively. We note that a fifth site involved in the trial (Bagamoyo, Tanzania(2)) was not represented among the samples considered here.

### Amplicon sequencing

To assay parasite genotypes we developed an approach using selective whole genome amplification (sWGA(10)) to enrich *P. falciparum* DNA, followed by a PCR amplicon approach based on the previously published NOMADS panel(11) to amplify specific genes of interest. In addition to the key target gene (*CSP*), we included primer pairs for nine other loci which have known associations with antimalarial resistance or encode prominent antigens: *CRT* (resistance to chloroquine and piperaquine), *DHFR* and *DPHS* (sulfadoxine-pyrimethamine), *K13* (Artemisinin compounds), *MDR1* (quinolines), *MSP2*, *PMI* and *PMIII* (piperaquine), and *AMA1* (**Figure 1** and **Supplementary Tables 2-4**).

For each sample we extracted DNA from the whole blood spot using a silica column-based method. We then made two key modifications to the protocol described by de Cesare et al.(11) to support sequencing on the PacBio platform (Methods). First, we incorporated sequence tags into the amplicon primers (based on M13 sequences), which we used to implement a two-stage PCR approach that incorporates sample barcodes and sequencing primers during the second reaction. Second, we then pooled the barcoded samples across 96-well plates for library preparation.

A total of 384 samples (including 11 technical repeats) were prepared using this approach and sequenced on the PacBio Sequel IIe platform at the University of Oxford, resulting in high-depth sequencing of 328 DNA samples from 319 individuals in this pilot experiment (**Figure 1**). Including the non-clinical repeat sample, the successfully sequenced samples comprise 320 infection timepoints from 319 (R21=110, Control=219) individuals, and eight technical repeats.

**Figure 2** shows the number of reads aligning to the target regions in each of these samples. With the exception of the non-clinical repeat sample, all remaining samples were sequenced to high coverage across all amplicons; the median read count across amplicons was >500 except at *MSP2* (median = 262; **Figure S1**). The *CSP* amplicon in particular was well covered with >500 reads in all successfully sequenced samples. We did note that coverage varied somewhat between amplicons and samples indicating that some additional balancing of amplicons and pools may be useful for future experiments. The non-clinical repeat sample (depicted with an asterisk in **Figure 2**) also had low read counts across some amplicons suggesting substantial amplification variability, as expected if input DNA fragment counts are low.

**Figure 2.**
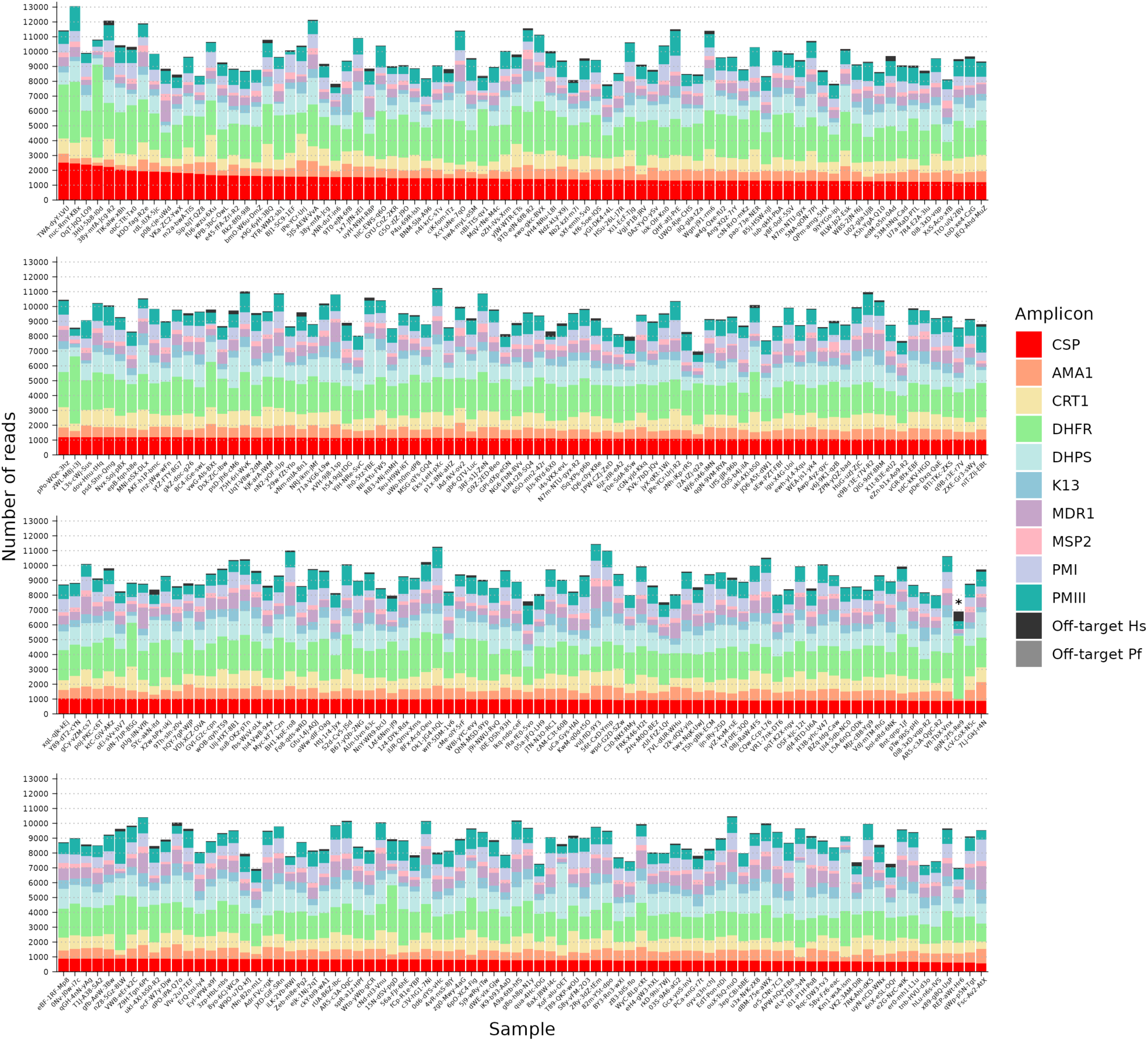
Amplicon balance in successfully sequenced samples from 319 clinical infections. The total number of PacBio reads (y-axis) is shown for each successfully sequenced sample (x-axis). Read counts are separated by amplicon target (colours), as computed by aligning reads to a reference sequence combining *P. falciparum* (Pf3D7) and representative assemblies of *P. ovale* (*P. ovale curtisi*, PocGH01), *P. malariae* (PmUG01), *P. knowlesi* (PKNH), *P. vivax* (*PvP01*), and the 1000 Genomes Project version of the GRCh38 human reference genome. Amplicon target coverage depends on various experimental factors including primer concentrations and binding efficiency, amplicon length (here ranging from approximately 3-4kb in length; detailed in **Supplementary Tables 2** and **4**). Asterisk denotes a single non-clinical infection timepoint (parasitaemia = 64 pRBCs/ul) taken one day post-treatment; all other samples met the study clinical infection definition (including microscopy positivity with >= 5,000 pRBCs/ul, and current or reported fever; Methods). Additional detail on amplicon balance and samples which failed sequencing is shown in **Figure S1**.

As outlined above, our samples also included a subset of 56 samples which were not successfully processed or where very little data was generated. In this context, we note that the sample set contains nine infections identified by microscopy as containing non-*P. falciparum* parasites (including four infections with *P. ovale* and five with *P. malariae*); six of these were among the above failed samples, as expected if they contain little *P. falciparum* DNA. The remainder of failures arose from miscellaneous technical failures, which is detailed in Methods. All failed samples will be re-sequenced in the planned full project.

### Calling full-length amino acid sequences from mono- and polyclonal infections

Generation of full-length sequence reads across the target genes provides an opportunity to predict the complete amino acid sequences present in these infections. However, two complications arise in achieving this. The first is that many infections are polyclonal for multiple *P. falciparum* genotypes, i.e. if they have arisen from bites from more than one infected mosquito, or from a single mosquito that itself carries multiple parasite genotypes. The second is that sequencing reads contain errors – in our data, both PCR amplification errors and sequencing errors are possible – so that it is not sufficient to directly call component haplotypes from the reads. Nevertheless, the accuracy of the PacBio HiFi method used here, and the large number of reads generated for each sample (**Figure 2**), suggests it should be possible to accurately call the infection composition from the reads. We developed a method to do this, focussing on predicted amino acid sequence variation at each target gene.

Our approach is fully described in Methods but in brief, we make the assumption that sequence errors are rare enough that the true infection haplotypes will be represented without error in a nontrivial proportion of the reads for each sample. For each gene and each sample, we therefore extract the portion of each read covering the gene using a set of predefined kmers (detailed in **Supplementary Table 5**), and use the most common haplotypes present in the reads as candidates for the true infection haplotypes. A process of read re-alignment is then used to infer the set of gene sequences in the sample and their relative frequencies. To allow the major source of error in PacBio sequence data, this method also compresses homopolymers internally (and will therefore not detect true differences between haplotypes that differ only in homopolymer tract length). For the analysis below we focus on haplotypes that are exactly supported by at least 10 reads in the sample and by at least 5% of informative reads. These criteria help avoid impact of low-frequency errors that can arise during PCR amplification.

This approach, which is implemented in a C++ program (“translatorator”; **Code Availability**), reports the inferred DNA sequences of the resulting candidates, their predicted amino acid sequence, and diagnostic information including the supporting read counts (**Figure S2**) for downstream analysis.

**Figure S3** shows a comparison of haplotypes inferred in nine pairs of repeat samples present in our data. These includes the non-clinical repeat sample, and eight samples that were obtained by re-amplifying and sequencing other samples. Most samples were inferred identically in both repeats albeit with some variation in the proportion of minor haplotypes. Some discrepancies were, however, observed in samples inferred to have low-frequency haplotypes in one repeat but as monoclonal in the other (e.g. Ipe-YCr-Urj at CSP; **Figure S3**). We also observed one apparently monoclonal infection in which the inferred *MDR1* haplotype differed by one base between repeats (ukl-4Xl-bS0). A notable additional discrepancy was that we also observed mixtures of haplotypes that differed only in lengths of repeats of homo-asparagine tracts (encoded by AAT repeats), which are known to mutate frequently during erythrocyte infection stages(12, 13).

A further natural sanity check of this method is whether the inferred sequences properly encode full-length amino acid sequences. Across the 8,797 called haplotypes we found two that were not a multiple of 3 in length (including one at *CSP*, mentioned further below) and four that had mutations causing apparent premature stop codons.

Overall, these results indicate that the haplotype calls generated this way are generally accurate, although by design of this process it will not identify rare infection components, and interpretation of specific sample haplotypes should perhaps be treated with caution.

### Study infections show high polyclonality

The availability of full-length haplotypes enables us to make lower-bound estimates of the polyclonality of infections by considering the number of haplotypes inferred for each sample at each locus. Initial investigation indicated that at *K13* and *MDR1*, apparent haplotype diversity was heightened due to the presence of variation in asparagine repeat lengths. After removing this variation, we found that for approximately half of samples (157 of 319) a single haplotype was inferred across all regions, indicating that the infection is monoclonal (or at least contains only low-frequency variation at these genes; **Figure 3A**). However, a substantial proportion of samples carried multiple haplotypes. The *MSP2*, *AMA1* and *CSP* gene regions were especially diverse; at each locus over 30% of samples had >1 inferred haplotype. The highest number of haplotypes was inferred for a single control-vaccinated sample (uyN-nCD-WNx, illustrated in **Figure 3B**) which carried six clearly different haplotypes at *CSP*, five at *MSP2* and four at *AMA1* and more than one haplotype at four of the other six loci in our panel.

**Figure 3.**
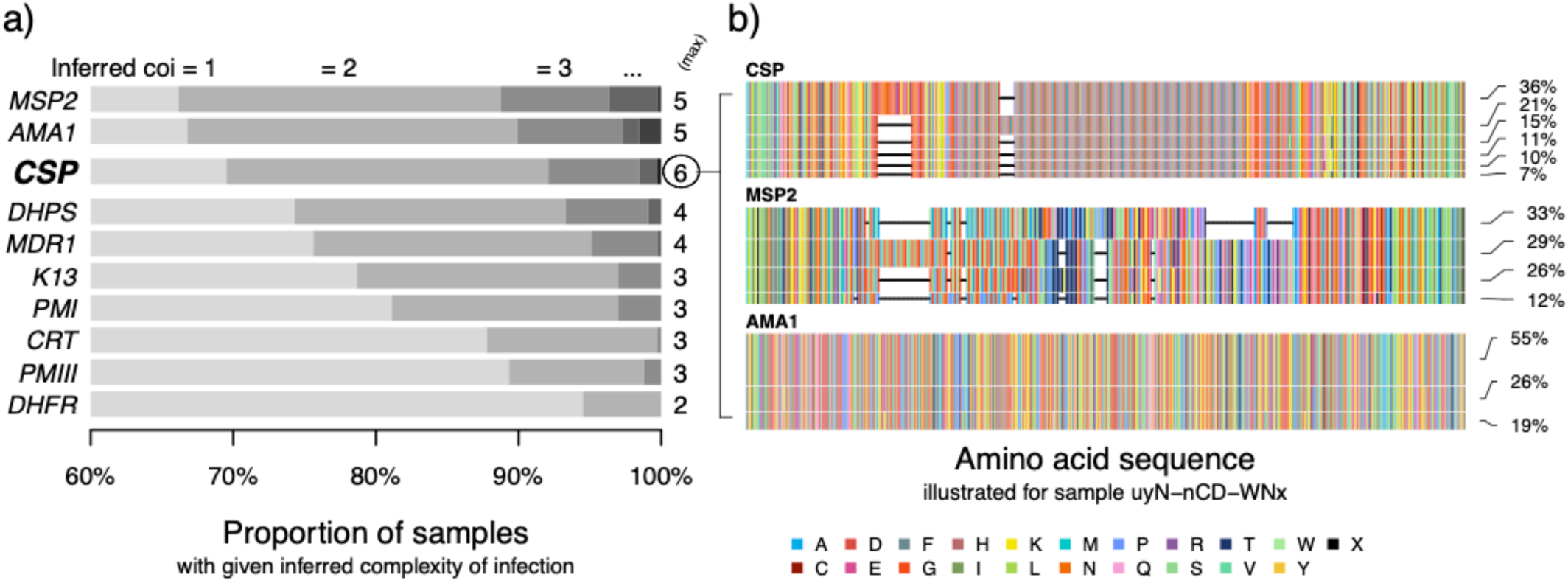
Detail of inferred polyclonal infections. **a)** the proportion of samples inferred to be monoclonal (light grey), or polyclonal with k>1 haplotypes (darker shades of grey) across the 10 amplicon regions (rows, ordered by polyclonal proportion). Results are shown after compressing all asparagine repeat segments (encoded by AAT or AAC codons). The maximum polyclonality observed across samples for each region is shown to the right. Sample uyN-nCD-WNx had the highest number of inferred haplotypes (6, observed at *CSP*) and is circled. **b)** Illustration of inferred amino acid haplotypes for uyN-nCD-WNx at the *CSP*, *MSP2* and *AMA1* loci. Each panels shows a multiple sequence alignment of the inferred amino acid sequences, with the inferred within-sample proportion shown to the right.

At *CSP*, just over 30% of samples (97 of 319) had evidence of more than one CSP amino acid haplotype in the read data, and across these samples the most common haplotype was inferred at between 36% and 92% of reads in the sample. In addition to the sample mentioned above, two samples (BNM-SJa-A96 and QhN-MSD-9Y) were inferred to contain four different CSP haplotypes; in both cases, these haplotypes differed in NANP/NVDP repeat length.

We noted weak evidence of association between higher multiplicity of infection (MOI) and higher numbers of infection episodes in the year since vaccination (e.g. *OR* = 1.33, 95% CI = 1.07 – 1.65 by proportional odds logistic regression of number of clinical infections against estimated MOI; *P* = 1.3 × 10^−2^; **Figure S4**). Illustrating this, the high polyclonality sample uyN-nCD-WNx was the seventh infection episode for this individual in this period, of which six reached the clinical infection definition, among the highest observed for any sample. MOI was also noticeably lower in R21 vaccine recipients compared to controls (rate ratio = 0.72, 95% CI = 0.56-0.94 by zero-truncated Poisson regression adjusted for study site and age group; *P* = 1.6 × 10^−2^; **Figure S5**). Sequencing of additional timepoints will be needed to determine whether some of the high MOI infections result from longstanding infections that are not fully cleared by treatment.

### The CSP amino acid sequence across 319 infections

The R21/Matrix-M vaccine comprises a fusion protein based on the C-terminal region and a section of an internal NANP repeat segment of the *CSP* gene(14). This is fused to a Hepatitis B surface antigen (HBsAg) virus-like particle (VLP) that enables relevant epitopes to be displayed on vaccine particles. The vaccine was designed using sequence from *P. falciparum* strain NF54, which is the progenitor of the 3D7 strain used to generate the reference sequence.

Analysis of available *P. falciparum* reference assemblies(15) and previous analyses(5) indicate that there is considerable variation in the amino acid sequence of CSP between parasite isolates. In particular, several segregating polymorphisms exist in the C-terminal segment, and there is substantial repeat length variation in the NANP/NVDP repeat (which has been observed to vary from 37 to 44 repeats in African populations(5) and larger in some non-African populations(16–18)). Variation is also observed at the N terminal, including a common insertion of 19 amino acids.

**Figure 4** depicts the inferred amino acid sequences for *CSP* in our data alongside that for 3D7. In total 449 amino acid sequences were observed across the 319 infections, but many were shared between samples for a total of 250 distinct sequences. The most commonly observed haplotype was observed 22 times (i.e. in just under 5% of sequences) and was 386 amino acids in length; meanwhile just over 40% of sequences were observed exactly once. Notably, none of the inferred haplotypes were identical to the 3D7 haplotype across the whole gene, and only a minority (35 of 450) were identical to the 3D7 / R21 vaccine sequence in the last 199 residues; of the remainder, most had around 6-9 differences (by edit distance) (**Figure S6**). As expected, most variation was observed within the central NANP/NVDP repeat (coloured blue and orange in **Figure 4**) and in the immunologically relevant Th2R and Th3R segments of the C-terminal segment, as well as at the N-terminal insertion(19) at positions 77-95 in the alignment (**Figure S7**). Notably, amongst common variant alleles seen in at least four haplotypes, all except four were observed in parasites from every study site in our data (**Figure S7**).

**Figure 4.**
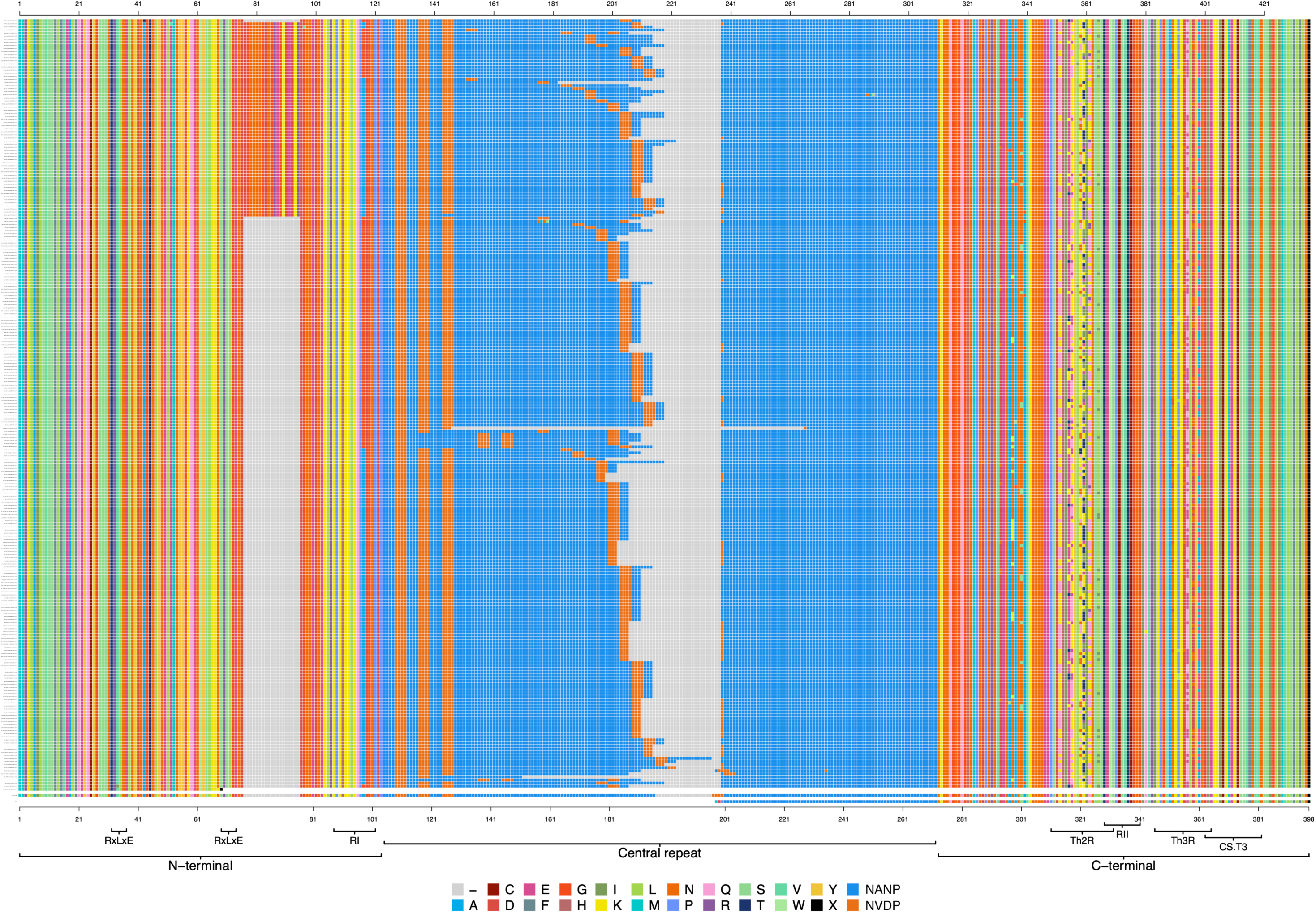
Full-length *CSP* amino acid haplotypes observed across N=319 infections. Rows show the distinct *CSP* amino acid sequences (colours; see legend at bottom) inferred across the study samples. In total 250 distinct haplotypes were observed, in addition to the 398-residue Pf3D7 reference sequence which is shown at the bottom along with the R21/Matrix-M sequence. Coordinates are given with respect to the Pf3D7 reference sequence gene (bottom axis) and with respect to the multiple alignment (top axis). The vaccine sequence is shown in the bottom row. Underneath, key regions of the *CSP* sequence are annotated including predicted PEXEL motifs (RxLxE), the RI and RII regions and the immunodominant Th2R and Th3R T cell epitope sequencing. Details of the sequences shown here are in **Supplementary Table 6**.

The number of combined NANP/NVDP repeats in major haplotypes in these samples varied from 37 to 46 repeats, with a slight trend towards higher repeat lengths in Kilifi (**Figure S8**). However, we also noted a small number of haplotypes in polyclonal samples that contained longer deletions or frameshifts, described below. Including the flanking peptides, the majority of repeat segments are of the form:

| 1 | 3 | 8-16 | 1 | 17-23 | 1 |
|---|---|---|---|---|---|
| NPDP | NANPNVDP | NANP | NVDP | NANP | NKNN |

where the numbers above the lines indicate the range of repeat counts observed.

Three samples were also inferred to have mixed infections in which one of the haplotypes has an especially short repeat segment falling outside these bounds (2Ry-3dZ-tEm, 33 repeats supported by 19% of reads; T89-QKP-wOU, 30 repeats supported by 11% of reads; yJW-WTR-ETs, 17 repeats supported by 16% of reads). This last example may be particularly notable as this repeat length is shorter than that encoded in the vaccine sequence (which has 18 full NANP repeats; bottom row of **Figure 4**).

Finally, we noted one sample (3HU−5b8−I0d) containing an apparent frameshift mutation causing an early premature stop codon (visible near the bottom of **Figure 4**). Inspection of reads for this sample indicates that this is caused by a one-base pair insertion in a homo-thymine stretch that is present in a majority of reads. A second insertion, downstream of the novel stop codon, is also present. *CSP* is regarded as essential for liver-stage infection and future resequencing will be needed to determine if this is caused by an amplification artifact.

### Comparison of population and within-infection *CSP* heterozygosity

Previous work has examined whether heterozygosity at specific amino acids observed in polyclonal infections is exceptionally high or low when compared to the population-averages(20, 21), potentially suggesting within-host selection effects. We examined this in our data using a simulation approach similar to previous work(20) (Methods and **Supplementary Table 7**). We notably found strong evidence for lower than expected within-infection heterozygosity at position 314 (observed vs. expected ratio = 0.7; empirical P = 0.0004; **Figure S9**), an effect which is however in the opposite direction to that reported in a previous study of RTS/S trial samples(20). High within-infection heterozygosity was also observed at position 317 (P=0.005). Other variants showed limited evidence for a within-infection effect (P > 0.01). We did note a trend toward high within-infection heterozygosity at positions 301 (compatible with Early et al.(20)) and 354 (compatible with Gilbert et al.(21), but in the opposite direction to Early et al.(20)) (**Figure S6**). We caution that, due to the relatively small sample size in this study, these analyses do not account for vaccination status and will need further replication.

### Comparison of CSP variation in control- and R21-vaccinated participants

The overall goal of the full study is to assess whether R21/Matrix-M vaccine efficacy varies between parasites with different genotypes. A suitably powered analysis of these effects will require data from the full trial (which will include all infection timepoints from at least two years post-vaccination). However, the pilot data generated here give the opportunity to make an initial assessment. We therefore summarised *CSP* variation in the R21 and control arms, across the four sites for which we have data (**Figure 5** and **Figures S6-8**), and undertook analyses to determine any preliminary evidence for differential infection rates.

**Figure 5.**
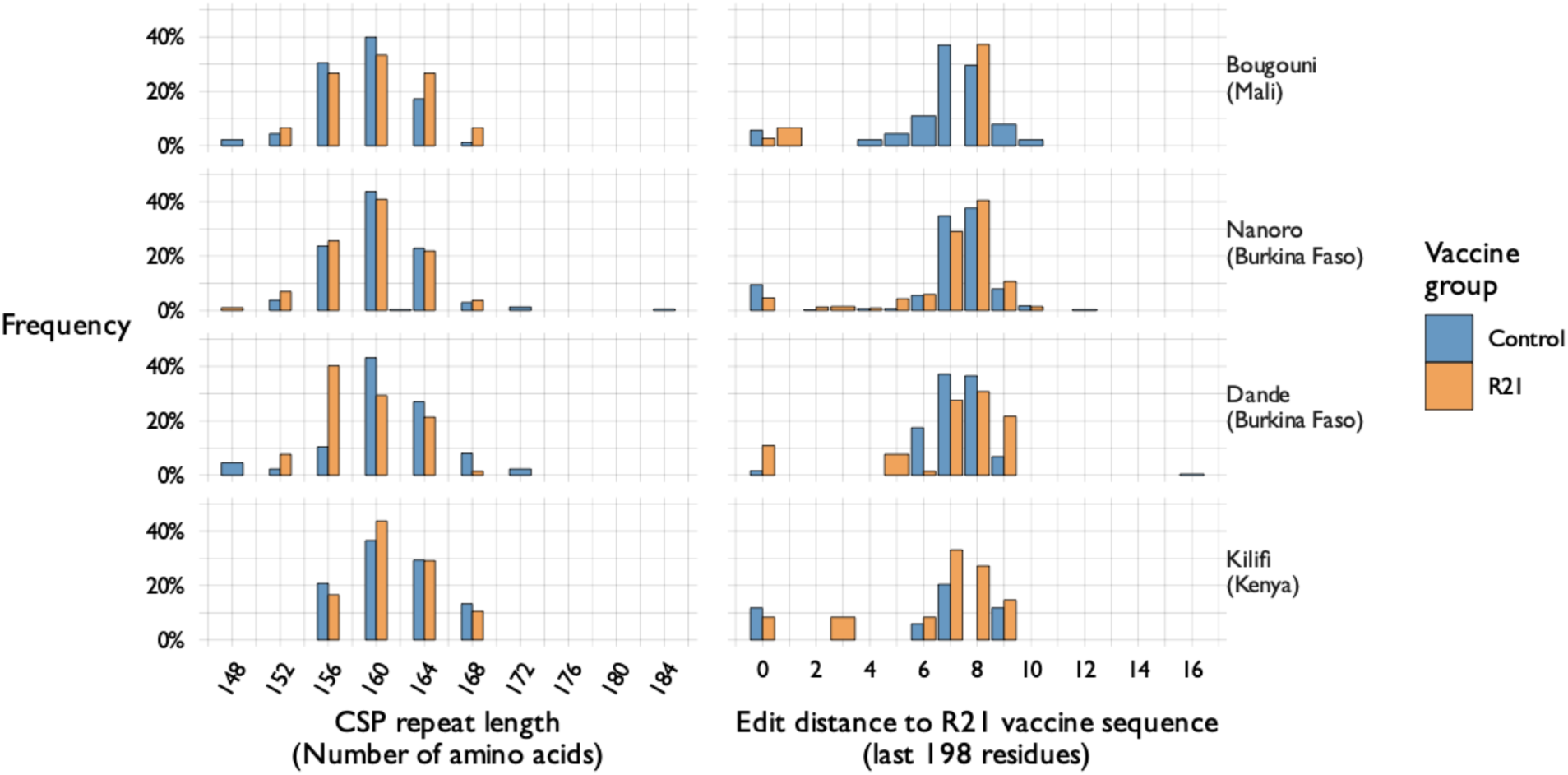
Distributions of CSP repeat length and distance to R21 vaccine sequence by trial site and vaccine group. For each amino acid repeat length (x-axis, left panel) or each observed edit distance to the R21 vaccine sequence in the last 198 residues (x-axis, right panel), the proportion of inferred *CSP* haplotypes with that repeat length (y-axis) is shown within each vaccine group (colours) and by site (rows). For polyclonal infections, proportions are accumulated based on within-sample proportions. To enable a visual comparison between vaccine groups the left panel x-axis is clipped to exclude three haplotypes with repeat lengths shorter than 140; both panels exclude one haplotype with an inferred premature stop codon referred to in the text.

Based on the observed haplotype variation we focussed on variation in the central repeat and the C-terminal segment, as well as across the length of the vaccine sequence which includes 18 NANP repeats and the whole C-terminal segment. We also assessed association with the other major variant observed in these samples, that is, the 19-residue insertion in the N-terminal segment.

The distributions of repeat length were similar between R21- and control-vaccinated arms at all four sites (**Figure 5** and **Supplementary Table 6**). The samples with the most extreme repeat lengths were all control-vaccinated; however, our data contain samples from a larger number of control-vaccinated than R21-vaccinated individuals (209 versus 110; **Figure 1**) so this is likely expected. A linear regression model test of variation of repeat length between vaccine arms also did not reveal any strong relationships (**Supplementary Table 8**; for simplicity, association analyses presented here are based on the most frequent haplotype observed in each sample and include study site as a covariate).

We also considered the evolutionary distances (measured as edit distance in the amino acid sequence) between study haplotypes and the full Pf3D7 C-terminal sequence, as well as to the last 198 residues of the full vaccine sequence which incorporate the C-terminal and 18 NANP repeats (**Figure 5** and **Supplementary Table 6**). In total 87 distinct haplotypes were observed across the last 198 residues and 83 across the C-terminal. 35 infections contained haplotypes matching the vaccine sequence exactly; these were distributed across control and R21 arms. However, the majority of samples differed by at least 5 amino acid changes from the reference haplotype (**Figure 5**). As for repeat length analysis, there were no statistically detectable differences in these distributions between vaccine arms (**Supplementary Table 8**).

Finally, we also examined the possibility of association between specific amino acid variants (identified as columns with variation in the sequence alignment in **Figure 5** and detailed further in **Figure S7**) and vaccine group. Results are presented in **Supplementary Table 8**. We again observed no strong evidence for association. Among alleles observed at least 5 times in these samples, the lowest P-value was observed for variation in the Th2R region which showed a nominally statistically significant trend toward higher frequency of the Pf3D7-matched allele in R21-vaccinated individuals (e.g. unadjusted P = 0.016 for effect of vaccine-matched Th2R versus non-reference alleles) and thus does not plausibly represent a vaccine escape effect. We did note some evidence that non-reference amino acids in the C-terminal position 299 (299A>D/G/S) were at higher frequency in the R21-vaccinated individuals; five samples had alternate amino acids at this position, four of which were in R21-vaccinated individuals (OR = 1.03; 95% CI = 1.00 - 1.06; unadjusted P = 0.03). These results, however, reflect tests across multiple alleles and very likely arise due to sampling variation. Inclusion of additional covariates (including episode and time since first vaccination) or including all haplotypes for each sample did not materially affect these estimates.

### Presence and haplotype structure of antimalarial drug-resistance associated variants

In addition to *CSP* analysis, we used the full-length haplotype data across *K13*, *DHFR*, *DHPS*, *CRT*, and *MDR1* to screen these samples for known antimalarial resistance mutations. The full panel of drug-resistance-associated variants we examined, as identified from literature sources, is shown in **Supplementary Table 9**. We briefly describe the results below; unless otherwise stated, only resistance-associated substitutions observed in at least one sample-derived haplotype are described below.

Mutations in *K13* are associated with partial artemisinin resistance. Across the 476 *K13* haplotypes called within this pilot set, we observed none carrying resistance-associated substitutions, including any of the 14 published WHO-validated resistance markers(22). This may be expected, as these samples date from 2021-2022, which is when the presence of artemisinin resistance was first reported in sub-Saharran populations, primarily in East Africa(23–26). Additionally, our pilot dataset did not include samples from Tanzania, where the presence of R561H has become well established(27). We did note, however, the presence of a non-synonymous mutation at codon 458, resulting in N458D, that co-locates with the resistance-associated N458Y substitution, in a single sample from Nanoro, Burkina Faso. To our knowledge, N458D has not been associated with artemisinin resistance, but has previously been observed in Ghana, Cameroon, and Côte d’Ivoire(28–30).

To examine resistance to other common antimalarials, we used our haplotype data to consider co-occurring resistance-associated substitutions across gene haplotypes. Within *DHFR*, pyrimethamine-resistance mutations occurred in structured combinations, with the triple-mutant N51I + C59R + S108N (IRNI) haplotype occurring in 278/337 (82%) of sample-derived haplotypes (**Figure 6**)(31). The rare I164L substitution was detected on two N51I + S108N haplotypes, both originating from Kilifi, Kenya, where I164L has previously been reported at low frequencies(32).

**Figure 6.**
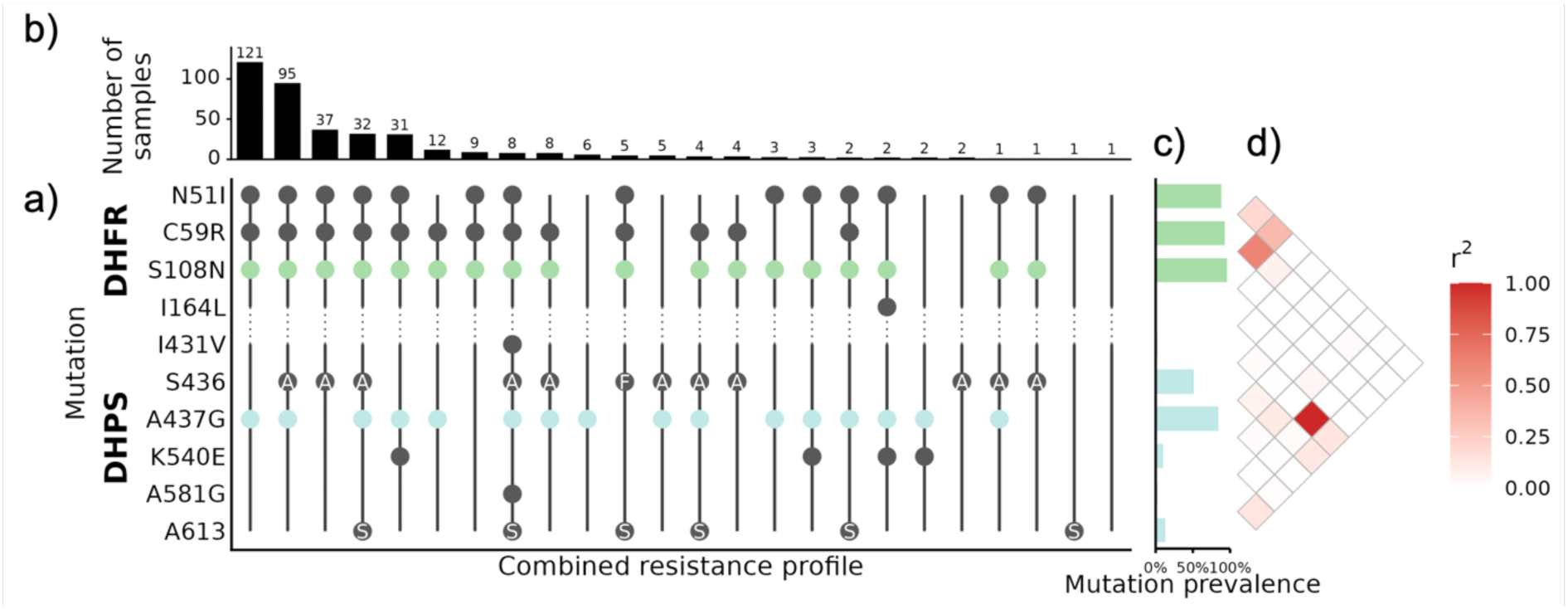
Combined sulfadoxine-pyrimethamine resistance haplotypes for *DHFR* and *DHPS*. **a)** Resistance associated substitutions comprising each *DHFR*-*DHPS* haplotype observed. Circles represent the presence of resistance-associated substitutions, with letters denoting the specific alternate residue at multi-allelic sites. The essential substitution for drug resistance is coloured in green (*DHFR*) or blue (*DHPS*). Samples containing multiple distinct resistance haplotypes at both loci were excluded from this figure, as phasing between amplicons could not be resolved (**Supplementary Table 10**). **b)** Number of samples in which each combined-resistance profile was observed. **c)** Marginal prevalence of each resistance-associated substitution across combined profile calls. **d)** Pairwise linkage disequilibrium (*r*^2^) between resistance-associated substitutions within and between *DHFR* and *DHPS*.

Across *DHPS*, sulfadoxine-resistance-associated substitutions showed marked geographic structuring consistent with that previously reported(33–35). K540E was strongly concentrated in Kilifi, Kenya, where it was detected in 28 of 29 haplotypes. In contrast, the A581G and I431V substitutions were detected exclusively in eight West African haplotypes, co-occurring on the same haplotypes in all cases. All eight additionally carried A437G and A613S, whilst retaining the wild-type K540 allele.

Motivated by the common use of sulfadoxine and pyrimethamine as partner drugs, we also examined combined gene haplotypes across the *DHFR* and *DHPS* loci (**Figure 6** and Methods). Little evidence of between-locus linkage disequilibrium was observed, and we noted that the predominant DHFR IRNI resistance haplotype occurred alongside several distinct DHPS haplotypes.

The *CRT* 72-76 CVIET haplotype, which confers strong resistance to chloroquine and is the target of a previous selective sweep(36), was observed in 45/354 (13%) sample-derived haplotypes, with prevalence highest in Bougouni, Mali (33%), followed by Dande, Burkina Faso (17%) and Nanoro, Burkina Faso (6%) (**Figure 7**)(37). CVIET was not detected among the 30 Kilifi haplotypes.

**Figure 7.**
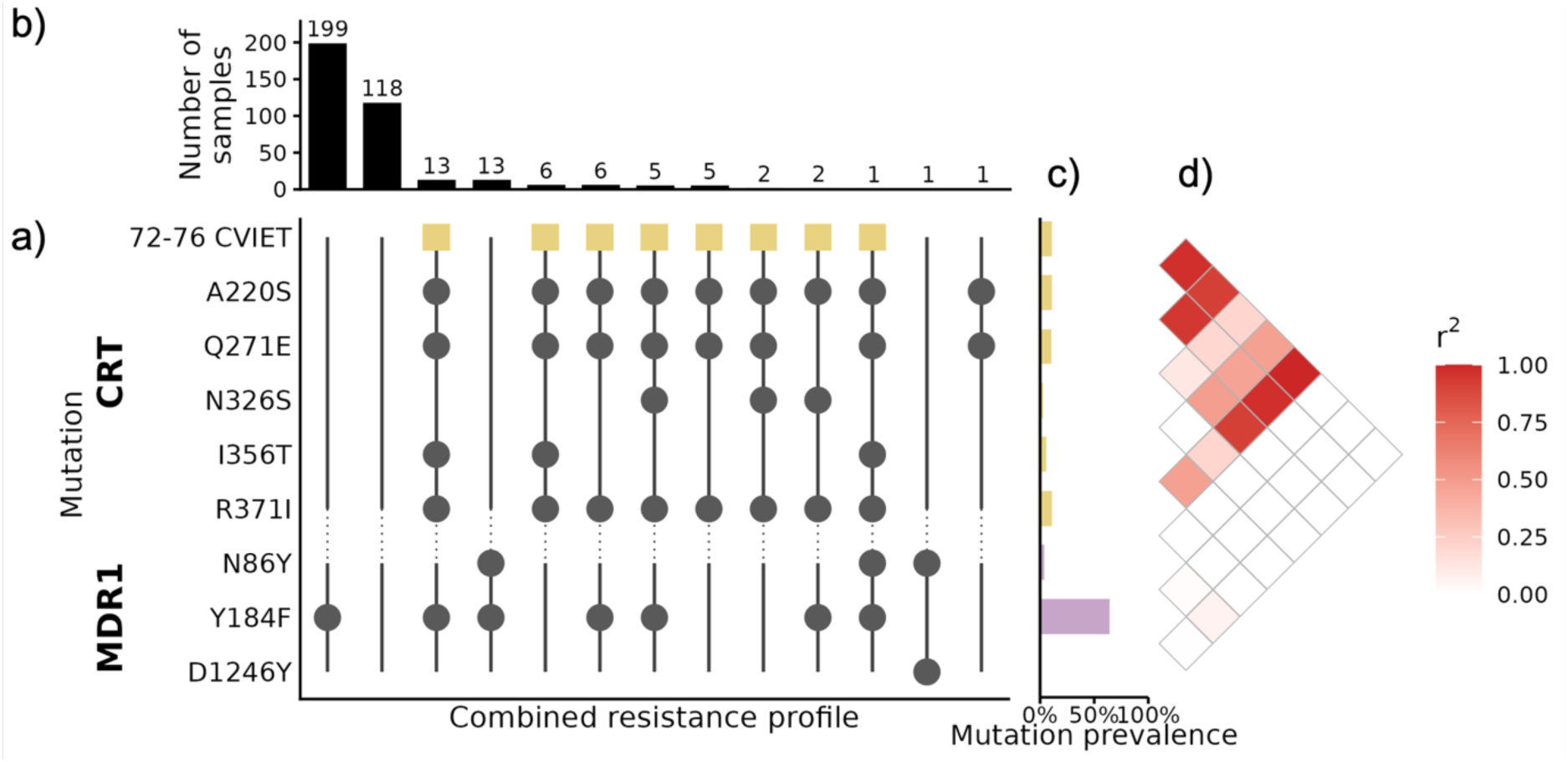
Combined chloroquine resistance haplotypes for *CRT* and *MDR1*. **a)** Resistance associated substitutions comprising each *CRT*-*MDR1* haplotype observed. Circles represent the presence of resistance-associated substitutions. The 72-76 CVIET/CVMNK haplotype is represented by a yellow square. Samples containing multiple distinct resistance haplotypes at both loci were excluded from this figure, as phasing between amplicons could not be resolved (**Supplementary Table 10**). **b)** Number of samples in which each combined-resistance profile was observed. **c)** Marginal prevalence of each resistance-associated substitution across combined profile calls. **d)** Pairwise linkage disequilibrium (*r*^2^) between resistance-associated substitutions within and between *CRT* and *MDR1*.

Additional *CRT* resistance-associated variation occurred almost exclusively on the CVIET background, reflected in high within-locus linkage disequilibrium (**Figure 7a**). An exception was a single CVMNK + A220S + Q271E haplotype observed in Nanoro, Burkina Faso. The CVIET haplotypes were also observed in combination with both wild-type and mutant haplotypes at the MDR1 locus, which has been associated with responses to multiple antimalarial drugs, including chloroquine, mefloquine, and lumefantrine(38).

Finally, we note that copy-number variation represents an additional mechanism of antimalarial drug resistance that is not captured by the amino-acid variant analysis presented here. Amplification of *MDR1* has been associated with altered susceptibility to lumefantrine and quinine, and amplification involving *PMIII* (and *PMII*, which is not included in this amplicon panel) has been associated with piperaquine resistance(39, 40). However, the current amplicon analysis was not designed to provide reliable gene copy-number estimates, and the presence of these resistance-associated amplifications could not be determined in the current data.

## Discussion

We have described the development and pilot results from a project to sequence clinical *P. falciparum* infections of children enrolled in the R21/Matrix-M phase III trial. The data represent infections from four study sites in three countries, arise from both R21-vaccinated and control-vaccinated children, and cover the *CSP* gene as well as nine other *P. falciparum* gene region amplicons. The results demonstrate that we are able to recover full-length amino acid haplotypes of these *P. falciparum* genes, use them to analyse mixture components in polyclonal samples, and conduct association analyses against trial metadata.

The data generated here provide a snapshot of the segregating genetic diversity circulating at the study sites during 2021-2022. It is notable that no infection in the study carried the same full-length haplotype as the reference strain Pf3D7, and more than 90% of observed haplotypes differed from Pf3D7 across the C-terminal segment alone. The fact that 3D7 is not representative of current population variation across the genome is well known, and is a potential concern for vaccine and therapeutic designs. On the other hand, the 3D7 C-terminal haplotype (which matches the vaccine design) is present at all study sites in our data, and the vaccine repeat sequence is also well within the range seen in most of the population samples analysed here.

Apart from *CSP*, our data are also informative about variation in other genes including several prominent antimalarial resistance loci which we have analysed in detail (**Figures 6-7** and **Supplementary Table 9**). Our panel also targets genes encoding blood stage antigens which show high levels of polymorphism, including *AMA1* and *MSP2*. Illustrating this, these infections clearly show the dimorphic structure and extensive repeat diversity(41) in the *MSP2* gene, which is also of current interest as a candidate vaccine target(42) (**Figure S10**).

The overall goal of the study is to determine whether parasite genetic variation might interfere with R21/Matrix-M vaccine efficacy. We emphasise that this pilot study does not currently provide evidence for this. However, the study is also not well powered to exclude this possibility and we are now turning our attention to analysis across a larger set of samples. In the first iteration we expect to analyse all clinical infection timepoints ascertained in the study within the first two years post-vaccination, amounting to approximately 4,500 infection episodes in the >1,900 study participants who experienced a clinical malaria episode during this period. We will implement similar methods to those described here and extend to full longitudinal sieve analysis(5) of infection genotype by vaccination status. In addition to the amplicon approach, we also expect to generate whole-genome sequencing data in a subset of samples to enable genome-wide estimation of strain mixtures, confirmatory haplotype analyses, and evaluation of potential effects outside of the *CSP* gene itself.

This will also enable analysis of species composition; the effects of interventions on non-*falciparum* species of malaria parasite including *P. malariae* and *P. ovale*, which occasionally occur in individuals observed in this study, is largely unknown. Overall, we expect this study to help establish the efficacy profile of R21/Matrix-M across *P. falciparum* subpopulations and, potentially, predict selection effects as the current vaccines are rolled out.

Finally, we note that the field is now moving toward new vaccine candidates including those that target blood and transmission stages, and combined vaccination approaches, and efforts are underway to develop vaccines for non-*falciparum* malaria species. The question of what drives variation in vaccine responses will be equally important for these vaccines. Methods that estimate the impact of both parasite and host genetic variation (including immunogenetic variation) on these vaccines will be increasingly needed to create granular estimates of efficacy and make accurate risk predictions across populations. Effective mechanisms for data sharing will also be needed to support comparative analyses. Consideration of these aspects at trial design stage is likely to be an important aspect of future studies, and will ultimately lead to improved understanding of natural and vaccine-induced immunity and the associated parasite response in global malaria-endemic populations.

## Methods

### Ethics and consent

Ethical approval for the R21/Matrix-M Phase III trial (VAC078 / NCT04704830) trial protocol was overseen by national ethics committess and regulatory authorities in each country, and by the University of Oxford and London School of Hygiene & Tropical Medicine Research Ethics Committees as detailed previously(2).

Informed consent was given in writing or by thumb print by a parent/guardian for all children participating in the trial, including specific consent for downstream genetic analysis.

### R21/Matrix-M trial methodology

The R21/Matrix-M Phase III trial was a randomized, controlled study conducted across five sites in Burkina Faso, Mali, Kenya, and Tanzania to evaluate the efficacy, safety, and immunogenicity of the R21/Matrix-M malaria vaccine in young children living in areas of moderate-to-high *P. falciparum* transmission. Children were randomized in a 2:1 ratio to receive R21/Matrix-M (referred to below as R21 for brevity) or a control rabies vaccine (Abhayrab) according to the trial protocol, with longitudinal clinical and parasitological surveillance performed to detect malaria infections and clinical endpoints(2). The trial started in April 2021.

For the primary case definition in the Phase III trial, clinical malaria was defined as the presence of an axillary temperature ≥ 37.5°C and/ or history of fever within the last 24 hours, and *P. falciparum* asexual parasitaemia >5000 parasites/µL as determined by blood slide. Data was collected by passive surveillance, i.e. sample ascertainment relied on families to bring children for treatment when symptomatic.

We focus here on the first 12 months of the study, specifically defined as the 365 day period following 14 days since completion of the primary series of vaccinations. Over this 12 month period, nontrivial vaccine efficacy of around 60-82% was observed(2) depending on site and age group (estimated as 1 minus the hazard ratio in a survival analysis of time to first infection, which assumes a constant vaccination effect over time). This therefore reflects a lower overall rate of infection among R21/Matrix-M vaccinees: just under 40% of the clinical infection episodes in this period (988 of 2,539) occurred in R21/Matrix-M vaccinees; this was higher in older children (18-36 months at first vaccination) and lower (around 33%) in 5-17 month old children.

Trial results(2) contributed to the overall decision to deploy the vaccine in children aged from 5 months living in regions of sub-Saharan Africa. Following these results, the trial was extended for a further two years resulting in a total of 48-months follow-up after the primary series of vaccinations. This enabled longer safety and efficacy follow-up as well as the assessment of the need for further booster vaccines to enhance protection. A follow-up in a subset of individuals, now at school age, is ongoing.

### Sample ascertainment

All samples from this study were collected as part of the Phase III trial described above. DBS samples were collected on Whatman 20mm filter paper according to a standardised protocol, including a blood draw and pipetting of 2 x 50µL drops onto one filter paper. All samples were labelled with sample identifier and date of collection. Blood slide measurement of parasitaemia was also taken on each visit, along with identification of malaria species. For each enrolled individual, malaria episodes were numbered starting with 1 for the first episode; return visits within two weeks (14 days) of each other were regarded as the same episode.

All samples were shipped to the Jenner Institute, University of Oxford, and stored at -80°C. Sample metadata was also stored by the Jenner Institute. For this pilot study we focussed on a subset of samples selected according to the following criteria:

- Participant age of 5-17 months at the time of first vaccination.
- Only clinical infections (defined as the presence of an axillary temperature ≥37.5°C and/ or history of fever within the last 24 hours, and slide-positive *P. falciparum* asexual parasitaemia >5,000 parasites/µL as measured by blood slides) were considered.
- Only a single infection timepoint was included for each participant (participants experiencing no infections in this period were disregarded). We specifically selected the last clinical infection within the 12-month period following the completion of the initial dosing regime for analysis.

For the work described here, DBS were transferred to the Centre for Human Genetics, University of Oxford in batches for DNA extraction and sequencing as described below.

In addition to the samples described above, 10 samples not meeting the definition of clinical samples above were used for assay and protocol development and initial sequence testing. A single sample from an independent cohort called the GAMCC case/control study, previously analysed using Illumina sequencing(43), was also included throughout the pipeline. These samples are referred to as ‘test’ samples below. Following initial sequencing, one of the test samples was determined to be a *P. malariae* infection.

### DBS processing and sample barcoding

To ensure accurate sample tracking we implemented a barcoding system using dual barcoded labels, and processed DBS as follows. First, each DBS was inspected to determine sample ID and date, and the corresponding label was identified. We used a barcode scanner to confirm the match between sample and label and then labelled a fresh 1.5mL snap-cap tube. The filter paper was inspected for any irregularities such as unusual colour or shape, and this was noted on the sample tracking spreadsheet. One of the two blood spots on the paper was then cut up into approximately 3mm diameter pieces using a clean pair of scissors and deposited into the tube, which was then sealed. For all samples we returned the second DBS to the freezer for ongoing storage / future use.

### DNA extraction

DNA was extracted from the cut DBS using the Qiagen DNA Micro Kit (Qiagen, Hilden, Germany, Cat no. 56304). This uses a silica-based spin column to isolate DNA from other cell lysates. The extraction protocol was based on the NOMADs protocol(11, 44) but to account for the large amount of material from each DBS, we altered the protocol as follows. First, 360µL Buffer ATL (tissue lysis buffer) and 40µL Proteinase K were added to each cut DBS, which were then incubated at 56°C and 900 rpm for 2 hours. 400µL Buffer AL (cell lysis buffer) was added to the samples, which underwent further incubation at 70°C and 900 rpm for 20 minutes.

Upon transfer of the lysate to the columns, the washes for isolation of the DNA followed the protocol according to the manufacturer’s recommendations. Prior to elution of DNA from the columns, the samples were incubated in the elution solution (nuclease-free water/AE buffer) at 56°C for 10 minutes to increase DNA yield.

For each set of extractions, a randomised selection of samples was quantified using the Qubit dsDNA high sensitivity (HS) assay (Thermo Fisher Scientific, Massachusetts, USA). An extracted DNA concentration of >1ng/µL was considered successful. During testing, DNA quality was also measured using the Tapestation (Agilent, California, USA).

In initial experiments, the extractions were eluted into 50µL of nuclease-free water. However, based on repeated measurements we noted that the DNA concentrations of some samples decreased rapidly on storage, with the effect particularly pronounced after freezing at -20°C overnight. Repeat experiments identified that this was a result of elution in the nuclease-free water, which was nontrivially acidic (pH 4.0-5.0), likely inducing degradation of the DNA by acid hydrolysis. To avoid this, we altered the protocol to elute into AE buffer (10 mM Tris-Cl, 0.5mM EDTA at pH 8.0) and stored the samples in the fridge prior to successful selective whole genome amplification.

### Selective whole genome amplification

All samples were processed through selective whole genome amplification (sWGA), which specifically amplifies A/T-rich DNA. sWGA reactions were performed as previously described(10). Two previously published primer sets, known as 10A and 6A, were used for the sWGA reactions (**Supplementary Table 3**). Briefly, 40ng of DNA for each sample was made up to a volume of 30µL using nuclease-free water. An sWGA master mix was added to the sample for a final volume of 50µL, containing 1 x Phi29 buffer (NEB, Massachusetts, USA, M0269L), 0.1mg/mL recombinant albumin (NEB, M0269L), 2.5μM of each primer pool, 1mM of dideoxynucleotide (ddNTP) mixture (NEB, N0447L), and 30 units of Phi29 enzyme (NEB, M0269L). For the reaction itself, the PCR machine (Tetrad 2, Thermocycler, Bio-Rad, California, USA) was set to run at 35°C for 5 minutes, 34°C for 10 minutes, 33°C for 15 minutes, 32°C for 20 minutes, 31°C for 30 minutes, 30°C for 16 hours, and finally at 65°C for 15 minutes, before cooling to 10°C.

Following this, the produced DNA mass was quantified via a Qubit broad range (BR) assay. Samples that did not produce an expected yield of >40 ng/uL were re-processed on another plate. Although it was noted that negative controls are prone to high DNA mass production due to primer amplification, such samples still had substantially lower concentrations than a successful sWGA amplification from presumed positive samples. Prior to DNA sequencing, the sWGA products were stored at 2-5°C for up to 96 hours, before being transferred to -20°C for longer term storage.

### Amplicon amplification

We used primer pairs developed by the NOMADs project(45) to amplify DNA regions containing the *CSP* gene from sWGA-amplified DNA, in a multiplex with several other genes of interest (**Supplementary Table 4**). All primers used in this project were supplied by Integrated DNA Technologies, Inc. The genes targeted were *CSP*, *CRT1*, *DHFR*, *DHPS*, *K13*, *MDR1*, *MSP2*, *PMI*, *PMIII*, and *AMA1*. KAPA HiFi HotStart (Roche, cat no. 07958935001) was used for all amplicon reactions.

All of the primer pairs except *MDR1* span regions that are larger than their target genes. In particular, in the Pf3D7 reference sequence, the *CSP* amplicon spans 3,107 bases extending approximately 45 bases upstream and 1,865 bases downstream of the gene.

For an initial test, we used the test samples and an initial set of 12 study samples, either using just the *CSP* primer pairs (for 6 samples) or the full NOMADs multiplex (16 samples) with the recommended primer concentrations to sequence these genes on the Sequel IIe platform (PacBio, California, USA). Libraries were created separately for each sample; for 15 libraries we also spiked in whole-genome sWGA product. We then assessed amplicon performance by plotting the proportion of reads aligning to each target region. This experiment demonstrated that all the above amplicon pairs produced sequenceable products and in reasonably balanced amounts, although the *K13*, *DHPS* and *DHFR* loci had the largest proportion of reads. For downstream experiments we adjusted the primer concentrations slightly, as described in **Supplementary Table 4**.

To process the high volume of samples considered here in a cost-effective and efficient manner, we then developed a method to avoid running full PacBio library prep for each sample. This method works by altering the amplicon process to implement a 2-stage PCR that incorporates sample barcodes in the 2nd stage PCR. To achieve this, the above PCR primers were first re-ordered including 5’ tag according to the PacBio M13 primer design. This adds modified DNA sequences (which were originally sourced from the M13 bacteriophage) to the 5’ of each primer (**Supplementary Table 4**).

We then used these primers with the PacBio M13 plate (cat no. 102-135-500) to generate barcoded amplicons for each sample. The PCR conditions were as follows: 1st round: 95°C for 3 min; 22 cycles of 98°C for 20s, 50°C for 15s, 60°C for 6 mins; 60°C for 10min and cool to 8°C. 2nd round: 95°C for 3 min; 2 cycles of 98°C for 20s, 60°C for 15s, 65°C for 6 minutes; 8 cycles of 98°C for 20s, 65°C for 1 min, 60°C for 6 minutes and cool to 8°C.

### Library preparation and sequencing

We applied the above method to 384 samples (including 372 infections, 11 technical repeats and the low-parasitaemia nonclinical repeat) derived from study infections as described in main text and **Figure 1**. Samples were processed in four batches of 96.

A subset of samples were used for full quality control (using the Agilent TapeStation platform) prior to sequence library preparation. All samples QC’d by this method appeared high-quality, with clear ’spikes’ of fragment length centring around 3-4kb as expected given the amplicon lengths. All 384 samples were then taken forward for library preparation but of these, two had low concentrations at the end of the library preparation steps and for simplicity we excluded these from the final pool. The resulting pool was sequenced on a single flowcell on the PacBio Sequel IIe platform at the Centre for Human Genetics.

### Processing and quality control analysis of sequence data

Read bases were called using the onboard SMRTLink software, using ccs version 6.3.0 with the --hd-finder option. This option detects reads with different consensus in the forward and reverse strand of the reads; these reads are identified by the addition of /fwd or /rev to the read name. We excluded these reads from our main analysis below.

We aligned all reads to a custom reference genome using minimap2. The reference was obtained by concatenating the *P. falciparum* (Pf3D7), *P. ovale* (*P. ovale curtisi*, PocGH01), *P. malariae* (PmUG01), *P. knowlesi* (PKNH), *P. vivax* (PvP01) genomes, together with the 1000 Genomes Project version of the GRCh38 human reference genome including alt contigs, decoy sequences, and EBV sequence(46) . Minimap2 was run using the `-x map-hifi` option. We also specified `-Y` (which specifies soft-clipping for supplementary alignments), `-L` (which writes long CIGAR strings in the CG tag), and `--eqx` (which annotates alignment mismatches with X in CIGAR strings).

To assess the sequence data across the 384 sequenced samples, we inspected the proportion of reads aligning either to each target region, off-target in the *P. falciparum* genome, or to the human genome (**Figure 2**). Numbers of reads aligning to each amplicon were calculated from alignment files using the python package ‘pysam’ (version 0.22.1). We found that most sequenced reads aligned to the amplicon regions (on average >98%). Where off-target reads were present, 53% on average were comprised of reads which aligned to the human genome. Where the presence of a mixed infection involving *P. malariae* or *P. ovale* parasites was noted, we did not observe any reads aligning to the genomes of these species. This is expected as both the whole-genome amplification and amplicons used here are designed to amplify the *P. falciparum* genome.

Of 382 successful sequencing reactions, 54 produced data where the CSP region was covered by less than 50 reads indicating that our method to ascertain this data for these samples had failed. Excluding these, the CSP region was covered by between 554-2521 reads (with an average of 1089 reads; **Figure S1**). Successfully sequenced samples also included 9 repeat samples (**Supplementary Table 1**) which we analyse further below.

### Gene haplotype calling in monoclonal and polyclonal infections

We implemented a bioinformatic process to call the full-length amino acid sequence from the PacBio amplicon reads for each sample as follows. Our approach is based on several assumptions:

i. Infections might be monoclonal (i.e. contain a single parasite genotype) or polyclonal (e.g. contain multiple genetically distinct parasites, as a result of co- or super-infection). Thus, each infection could contain more than one allele of the focus gene, and the method must attempt to infer this.
ii. The data will primarily consist of full length reads, i.e. covering the full region between amplicon primer sequences for each target gene. However, amplification and PCR errors could occur so that off-target and/or chimeric reads might also occur.
iii. Reads are expected to have high accuracy, but sequencing errors can also occur. The major mode of sequencing error is expected to be incorrect calling of the length of longer homopolymer tracts. However, PCR errors may also occur and could in some circumstances generate erroneous fragments at nontrivial frequencies, e.g. if occurring early in amplification process.
iv. All analysable samples have a high number of reads (e.g. at least one hundred) aligning to each amplicon region.

The method presented here focusses specifically on the protein-coding sequence of each target gene. To do this, for each target gene we first identified a set of DNA kmers delineating the coding sequence regions of the gene and used these to extract gene coding regions from each read. Kmers were defined based on the Pf3D7_v3 reference genome annotation but we used a custom approach to allow for possible polymorphism in the kmer regions, including that observed between available *P. falciparum* genome assemblies(15, 43, 47) and/or in the MalariaGEN Pf7 population genetic resources(48). We allowed for mutations at these sites by including IUPAC ambiguity codes in the kmers. We also adjusted kmers to avoid impact of homopolymer length errors in the reads, including allowing for variation in the length of long homopolymers internal to some kmers, and inclusion of bases outside CDS regions in some kmers to anchor their location in the reads. Finally, our kmer alignment process allows for up to one additional single-base mismatch when searching for kmers in the reads. The kmers used for this work are detailed in **Supplementary Table 5**.

For each read aligning to the target amplicon region, we then searched for linear chains of kmers (matching in either orientation in the sequence) in the read. If a unique such chain was observed we extracted the relevant portion of the read for downstream analysis; reads with no matching chain or with more than one chain were discarded as uninformative. **Figure S2** details the proportion of informative reads observed for each sample at each amplicon.

The *CRT* and *DHPS* genes each have multiple coding exons, and therefore the portion of each read extracted by this method reflects a set of non-overlapping segments in the original read. In particular, *CRT* has 13 coding exons and this results in a lower overall proportion of reads regarded as informative in our analysis below (**Figure S2**); future adjustments to this process could improve the number of reads used at this locus.

The process above is thus customised for our amplicon approach and aims to extract only the bases exactly corresponding to coding sequence from each read. Some of the extracted sequences nevertheless still contain errors and so a further approach is needed to infer the true haplotypes present in each sample. To do this, we first removed the major source of PacBio errors by compressing all homopolymer tracts in the reads to single bases. For multi-exon genes, homopolymer compression was carried out within each exon separately. We then identified candidate haplotypes as those observed at nontrivial frequency in the sample (specifically those present in at least 5% of compressed reads in the sample). To ensure interpretability we also required that at least one uncompressed sequence supporting the candidate was observed in at least 10 reads. A stochastic search algorithm was then used to further refine this list of candidates.

We estimated the within-sample frequency of component haplotypes by re-aligning all the homopolymer-compressed read segment to the inferred candidate haplotypes using the WFA2 library(49). Reads were assigned to the best-matching candidate if the alignment identity was at least 95%; any read not meeting this threshold or aligning to more than one candidate was discarded. We estimated within-sample frequency by counting the reads assigned to each candidate. Finally, we output the DNA sequence of the resulting candidates, their inferred within-sample proportions and their predicted amino acid sequence for downstream analysis. Diagnostic information including the supporting read counts for each amplicon are shown in **Figure S2**.

A C++ program (“translatorator’) implementing this approach is available as detailed in **Code Availability.**

### Repeat sample analysis

To help assess the accuracy of our process, our sequencing included 9 infections sequenced in repeat as detailed in **Supplementary Table 1**. To assess consistency of our approach across repeats, we plotted the inferred proportions of component haplotypes for each pair of repeat samples using colours to indicate matching or non-matching haplotypes (**Figure S3**). Most infections were monoclonal at each locus, but a subset had evidence of polyclonality. We noted differences between repeat samples as:

• Presence / absence of an inferred low-frequency haplotype differing between some pairs. These plausibly arise due to changes in haplotype frequency during amplification steps.

• Similar but non-identical haplotypes observed in repeats differing by individual SNPs, which again are plausibly due to PCR artifacts.

• We also noted that some haplotypes differed in stretches of triplets of AAT repeats, which encode asparagine residues. These are known to mutate at high rates during erythrocyte stage infections(13, 50), so that it is plausible that these reflect random amplification effects on an underlying heterogeneous set of haplotypes. However, it is also possible that effects of this type could be caused by artifacts of sWGA or the KAPA HiFi-based amplicon approach we have used here.

### Polyclonal infection analysis

To inspect haplotype inference in individual polyclonal infections, we used MAFFT to create a multiple sequence alignment of the inferred haplotypes (either using the DNA sequences or inferred translated amino acid sequences). Results for one sample are shown in **Figure 3**. To estimate the overall multiplicity of infection (MOI) for each sample, we used the maximum number of haplotypes observed for the sample across all amplicons.

We assessed the relationship between inferred MOI and the number of episodes observed per sample (**Figure S4**) using a proportional odds logistic regression model, using the number of clinical episodes observed for each individual within 365 days of day 14 post-vaccination as the outcome variable, and using either MOI and an indicator of study site (results reported in main text), or MOI and study site, age at vaccination (measured in months), and vaccination status (R21/control) as predictors, with similar results.

To assess the relationship between inferred MOI and vaccination status, we fit a zero-truncated Poisson regression model using inferred MOI as the outcome variable, and vaccination status, indicator of study site, and age group at vaccination (5-12 months and 13-24 months as defined in the original Phase III trial) as predictors, using the VGAM(51) package in R. Model fit was assessed by comparing observed and model-predicted MOI frequencies separately for vaccinated and unvaccinated individuals (**Figure S5**).

### Calling CSP protein sequence variation between samples

We constructed a multiple sequence alignment of CSP allotypes across all the samples and available *P. falciparum* reference genome assemblies (**Figure 3**). We used this to identify regions of each sequence, to call repeat sequence content, and to identify between-sample variation in the N-terminal and C-terminal regions of the gene (**Supplementary Table 6**).

### Within-infection heterozygosity analysis

To assess evidence for unusual patterns of within-infection heterozygosity, we replicated a simulation approach used in a previous study(20). We focussed on the 97 infections inferred as polyclonal at the *CSP* locus in our data across the four study sites. For each site of variation in amino acids (**Figure S9**), we then computed the average heterozygosity of the site. Specifically, if *K* alleles were observed at the site in total, and these have within-sample frequencies *p*_1_, *p*_2_, ⋯ , *p_K_* as inferred from the haplotypes in the infection, we computed the heterozygosity as:

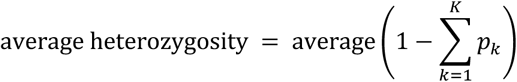

where the average is over the 97 polyclonal infections.

To compare this to an expectation, we then simulated sets of 97 infections matching the polyclonality and haplotype frequency composition of the observed infections. Haplotypes in each simulated infection were drawn at random from the haplotypes observed across all samples in that study site, weighted by the overall frequency of the haplotype. In total 10,000 simulations were performed generating 10,000 sets of 97 polyclonal infections for comparison; we then computed the average heterozygosity across each simulated set. The distribution of heterozygosity for simulated datasets at three specific amino acid sites is shown in **Figure S9**. Finally, we computed an ‘effect size’ as

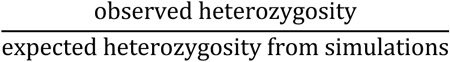

A standard error, 95% confidence interval and P-value were then computed based on the distribution of simulated heterozygosities.

### Haplotype vs. vaccine group association analysis

We used linear and logistic regression models to assess association between vaccine group and CSP haplotype variation as detailed in **Supplementary Table 8**. For each CSP variation measure, the vaccine group (R21-vaccinated, compared to control-vaccinated) was included as the predictor and the CSP variation was included as the outcome. For the analysis presented in main text we included an indicator of the site as a covariate to control for between-site variation in CSP sequence, and we restricted to the most common haplotype within each sample. Analyses including additional covariates (episode number and time from first vaccination to the episode) were also carried out with similar results (data not shown).

### Screening for drug-resistance associated substitutions

A predefined panel of amino-acid substitutions associated with antimalarial drug resistance was compiled for K13, DHFR, DHPS, CRT, and MDR1 (**Supplementary Table 9**). For each aligned long-read haplotype, the amino acid observed at each predefined position was extracted and compared with the corresponding reference and resistance-associated alternative residue(s). Calls matching the reference residue were classified as reference (ref), those matching a predefined resistance-associated residue as alternative (alt) and unexpected residues or gapped positions as other. At sites where multiple alternative alleles were possible, the exact amino-acid substitution was preserved when constructing resistance haplotypes. Haplotypes producing other calls were inspected separately for potential sequence or alignment artefacts prior to downstream analysis. Two MDR1 haplotypes were excluded from downstream analysis: one was extensively truncated across the amplicon (nN2-y0F-lUp-116/732-15.8%-12) and the other non-callable for residue 1246 (hwA-myL-oSM-233/539-43.2%-15). This resulted in a remaining 566 MDR1 haplotypes for analysis.

For each gene, resistance-associated substitutions occurring on the same long-read amplicon haplotype were combined to generate within-gene resistance haplotypes. Haplotypes containing no predefined resistance-associated substitutions were defined as wild type (WT). For CRT, residues 72-76 were concatenated to reconstruct the well-established 5 amino-acid CRT haplotype. Within this dataset, haplotypes segregated as either CVIET or CVMNK; M74I, N75E and K76T were therefore represented as the 72-76 CVIET/CVMNK haplotype in downstream analysis, while other CRT mutations were retained individually.

For the analysis of resistance profiles across loci associated with resistance to the same antimalarial treatment, within-gene haplotypes for DHFR/DHPS and CRT/MDR1 were paired. Samples containing a single resistance haplotype at both loci were directly paired. Where multiple distinct resistance haplotypes were observed at one locus but only one at the other, each haplotype was paired with the single haplotype at the corresponding locus. Samples containing multiple resistance haplotypes at both loci were excluded from combined-profile analysis (**Supplementary Table 10**).

For mutation-frequency and LD analysis, resistance-associated positions were encoded as binary variables representing the presence (1) or absence (0) of any predefined alternative-amino acid residue (or haplotype in the case of CRT residues 72-76) within the inferred combined resistance profiles. Marginal frequencies were calculated as the proportion of profile calls carrying each resistance residue/haplotype. Pairwise LD was calculated as the squared Pearson correlation between binary mutation states and summarised as *r*^2^. Within- and between-gene pairwise *r*^2^ values were visualised alongside the corresponding resistance-profile mutation matrices.

## Data Availability

Parasite genomic sequencing read data from this study will be made available via the European Nucleotide Archive (Study accession PRJEB130663).

## Code Availability

The translatorator haplotype calling software is available via github (https://github.com/gavinband/iorek).

## Acknowledgements

This study uses samples from the VAC078 clinical trial (https://clinicaltrials.gov/study/NCT04704830). It was financially supported by Serum Institute of India PVT Ltd. (SII), who are the primary manufacturers of the R21/Matrix-M vaccine. SII played no role in the design, implementation, interpretation or reporting of the study.

L.J.J. was supported by SII. G.B. and L.J.J. were supported by Wellcome ([304926/Z/23/Z](https://www.sciencedirect.com/unsupported_browser#gs1)). A.J.M was supported by the Chinese Academy of Medical Sciences (CAMS) Innovation Fund for Medical Science (CIFMS), China (2024-I2M-2-001-1).

We are grateful to the participants, families and caregivers involved in this study, and the local Health District authorities and hospitals involved in the trial, and to all the research staff at Ifakara Health Institute, Bagamoyo; Institut des Sciences et Techniques, Dande; Kenyan Medical Research Institute–Centre for Geographical Medical Research–Coast, Kilifi; Malaria Research and Training Centre, Bamako; Unité de Recherche Clinique de Nanoro, Nanoro who contributed to the clinical trial.

## Author contributions

Conceptualization: A.J.F., A.V.S.H., A.J.M., M.S.D., G.B.

Methodology: L.J.J., A.J.F., J.D., K.R., Y.W., A.C., J.H., P.P., A.J.M., G.B.,

Investigation: L.J.J., A.J.F., J.D., K.R., G.R., A.C., P.T-S.

Resources: K.R., A.F., S.W., B.H., A.B., E.B., A.D., M.H., A.O., J-B.O., H.M.N., H.T., L.S.

Software: G.B.

Formal analysis and visualisation: L.J.J., A.J.F., T.R., G.B.

Supervision: L.S., A.J.M., M.S.D., G.B.

Project administration: S.W.

Writing - original draft: L.J.J., A.J.F., G.B.

Writing - Review & Editing: L.J.J., A.J.M., G.B. and all authors

Funding acquisition: A.J.F., A.V.S.H., A.J.M., M.S.D., G.B.

