## Supplementary Figures (S1-10) for "Pilot long-read amplicon sequencing of clinical malaria infections from the R21/Matrix-M Phase III vaccine trial"

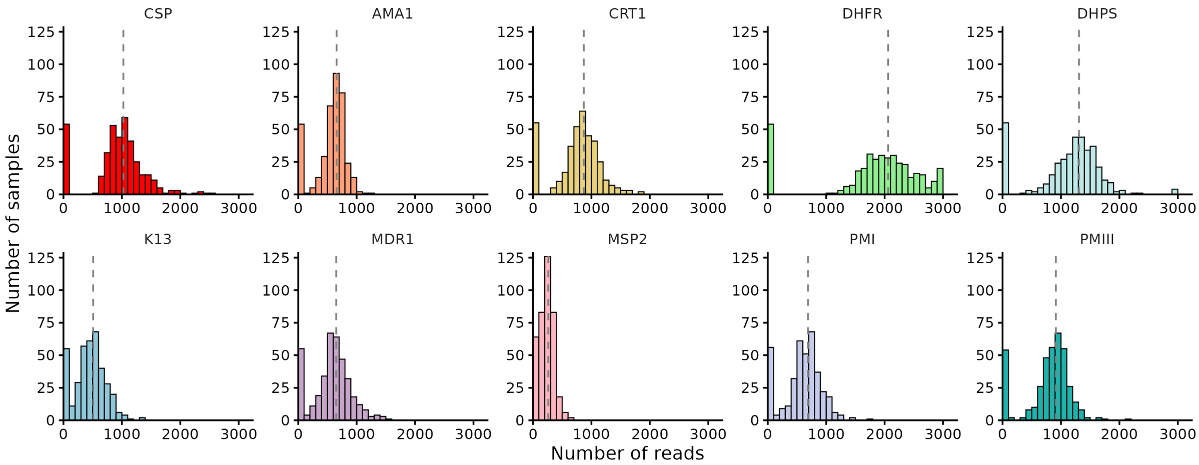

**Figure S 1 Distribution of sequencing read depths across amplicon regions.**

Histograms showing the distribution of read across all samples for each amplicon region, with reads grouped into bins of 100. Read counts >3000 were included in the last bin. Dashed vertical lines show the overall median number of reads calculated for samples considered successfully sequenced.

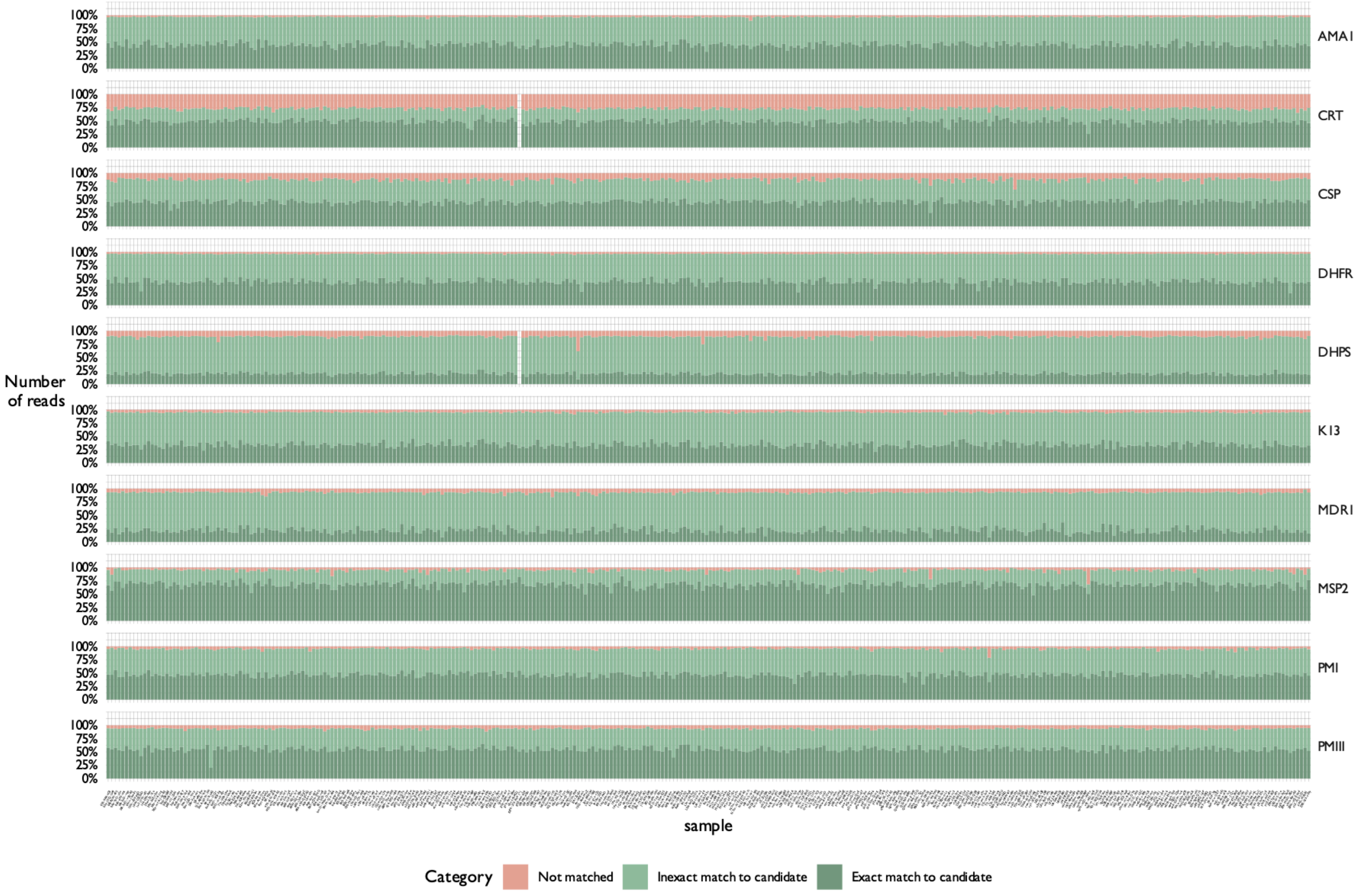

**Figure S 2 Detail of read count support for inferred haplotypes.**

Green, dark green and red bars show the proportion of reads for each sample (columns) that exactly matched an inferred haplotype (dark green), inexactly matched an inferred haplotype based on best alignment (lighter green), or were not matched to any inferred haplotype (red) for each amplicon region in our panel (rows). Unmatched reads include those that either did not contain the gene-identifying kmers (to within 1 basepair mutation), or had less than 95% identity to any candidate haplotype after homopolymer compression. The low-parasitaemia, nonclinical repeat sample (ggN-2f5-Re9) had no reads at the *CRT* and *DHPS* regions.

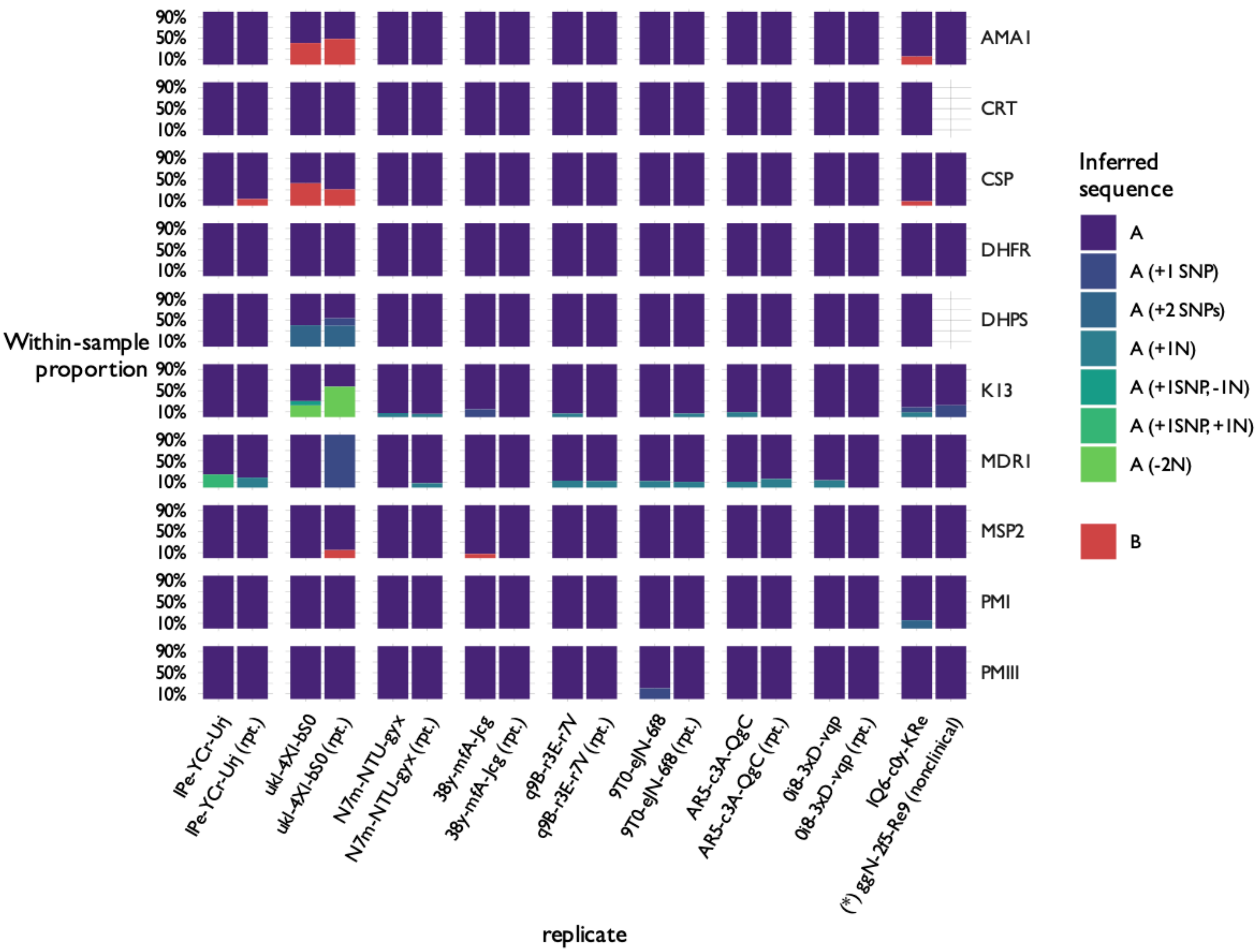

**Figure S 3 Comparison of inferred haplotypes across repeat samples.**

For each pair of samples repeated in our study (x axis), the inferred haplotype composition of the sample is shown at each target gene (rows). Colours depict the haplotypes inferred in the sample, with blue-yellow shades indicating haplotypes identical to, or similar to the most common haplotype inferred, and 'B' the second most common haplotype inferred. For visualisation purposes we classified haplotypes as similar if the edit distance was $\leq 6$, and for these classified the form of similarity as involving 1 or 2 SNPs and/or alteration of the length of an asparagine repeat sequence (N) as shown in the legend. All haplotype A/B pairs differed by at least 50 edits.

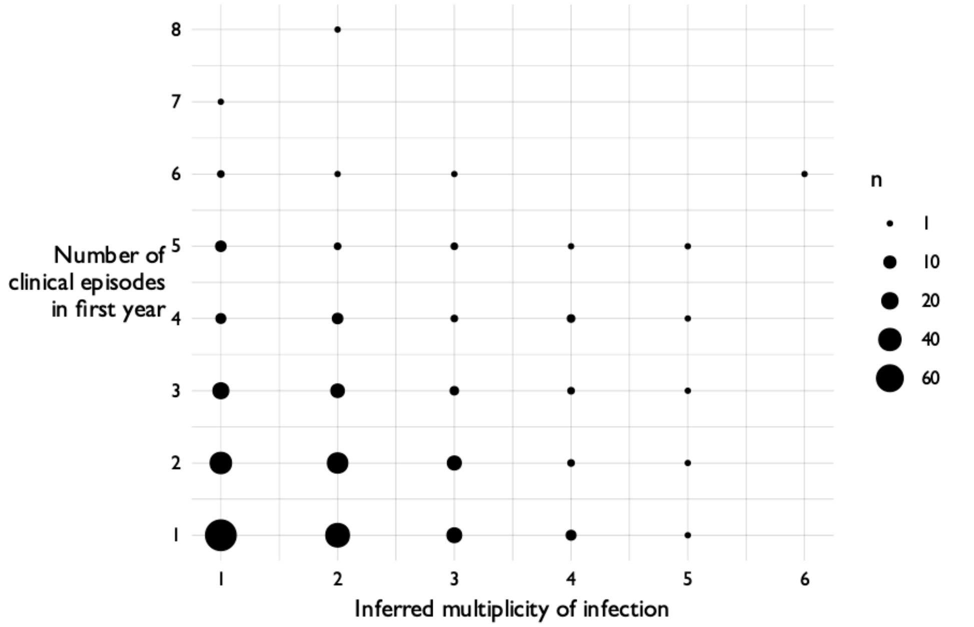

**Figure S 4 Comparison of inferred multiplicity of infection and number of clinical episodes across 319 infections**

Points show the number of samples (point size) having the given inferred multiplicity of infection (x axis) and the number of clinical episodes observed in the 12-month period following 14 days after the 3$^{rd}$ vaccination. Multiplicity of infection was taken as the maximum number of haplotypes inferred for the sample, over the 10 target genes. Distributions of multiplicity are shown in **Figure 4**.

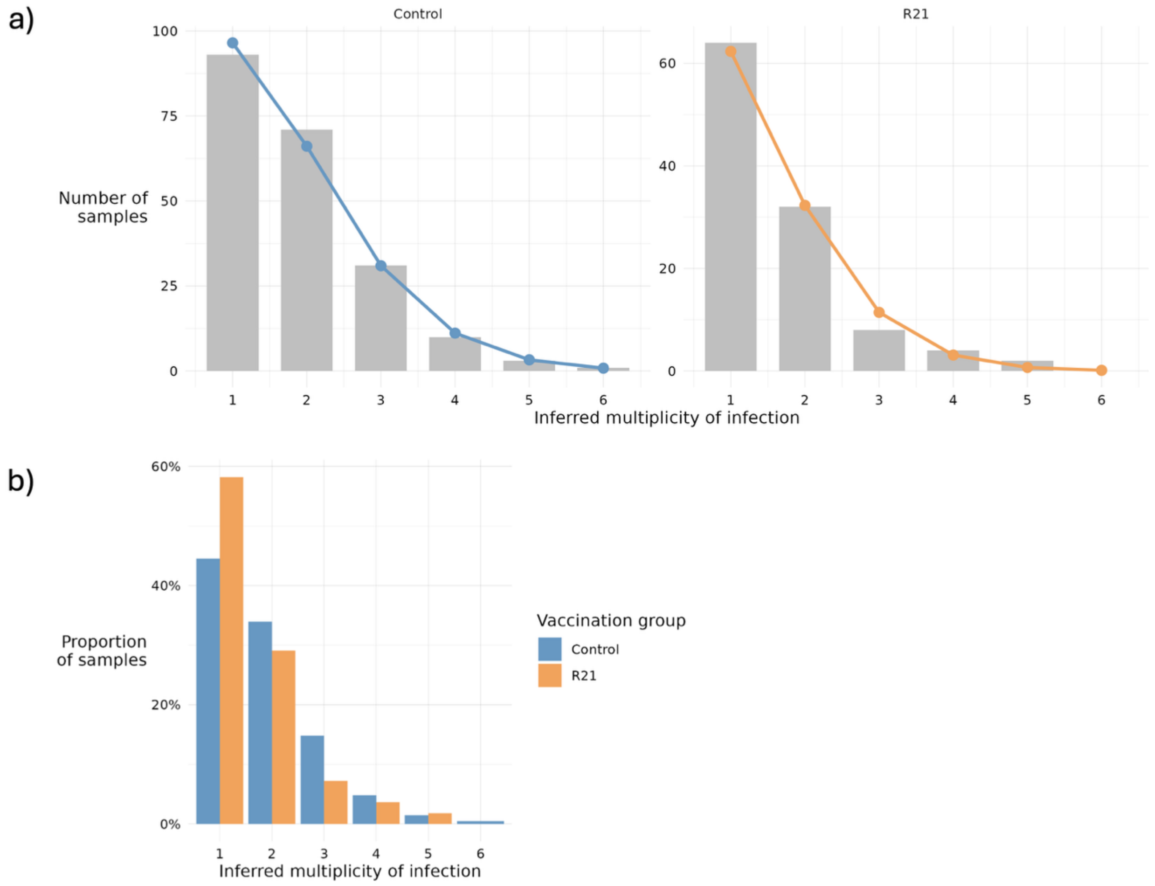

**Figure S 5 Comparison of inferred multiplicity of infection and vaccination status across 319 infections**

**a)** Observed number of R21 and Control (y-axis) samples having the given inferred multiplicity of infection (x-axis). The expected frequencies from a zero-truncated Poisson regression model adjusted for vaccination group, study site, and age group, is shown separately for R21 (orange) and Control (blue) samples. **b)** Proportion of R21 (orange) and Control (blue) samples (y-axis) having the given inferred multiplicity of infection (x-axis).

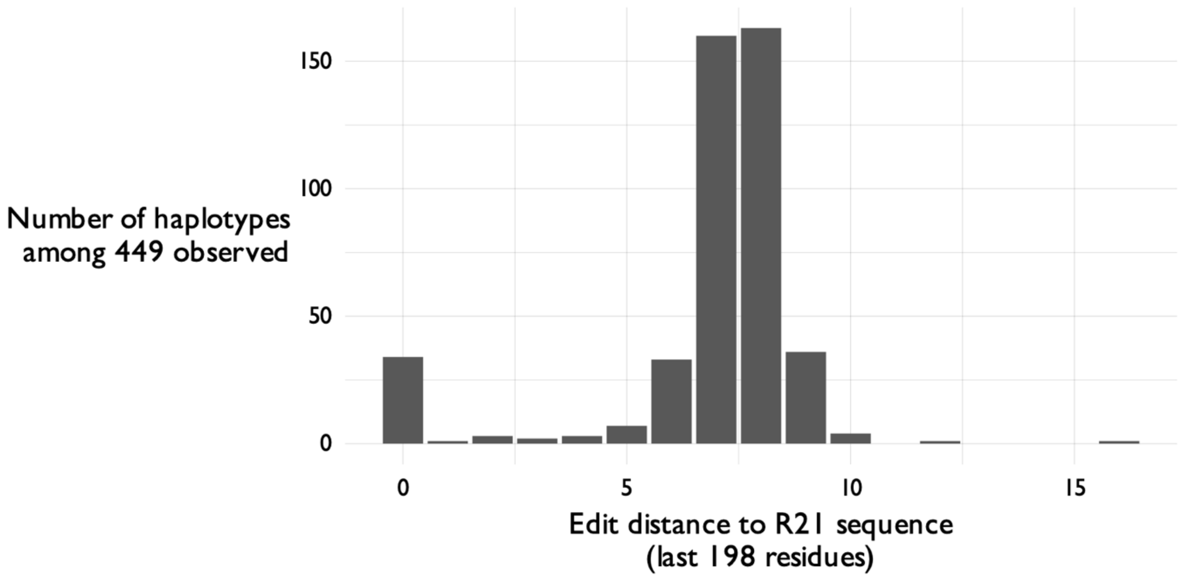

**Figure S 6 Haplotype distance to R21 vaccine sequence across inferred haplotypes.**

Histogram shows the edit distance between each CSP amino acid haplotyped inferred in our study and the R21 / Pf3D7 vaccine sequence. We note that the R21 sequence has length 201 and initiates with Methionine, Aspartate and Proline residues that are not present in any parasite sequence, as shown in **Figure 5**; the comparison above is therefore across the last 198 residues.

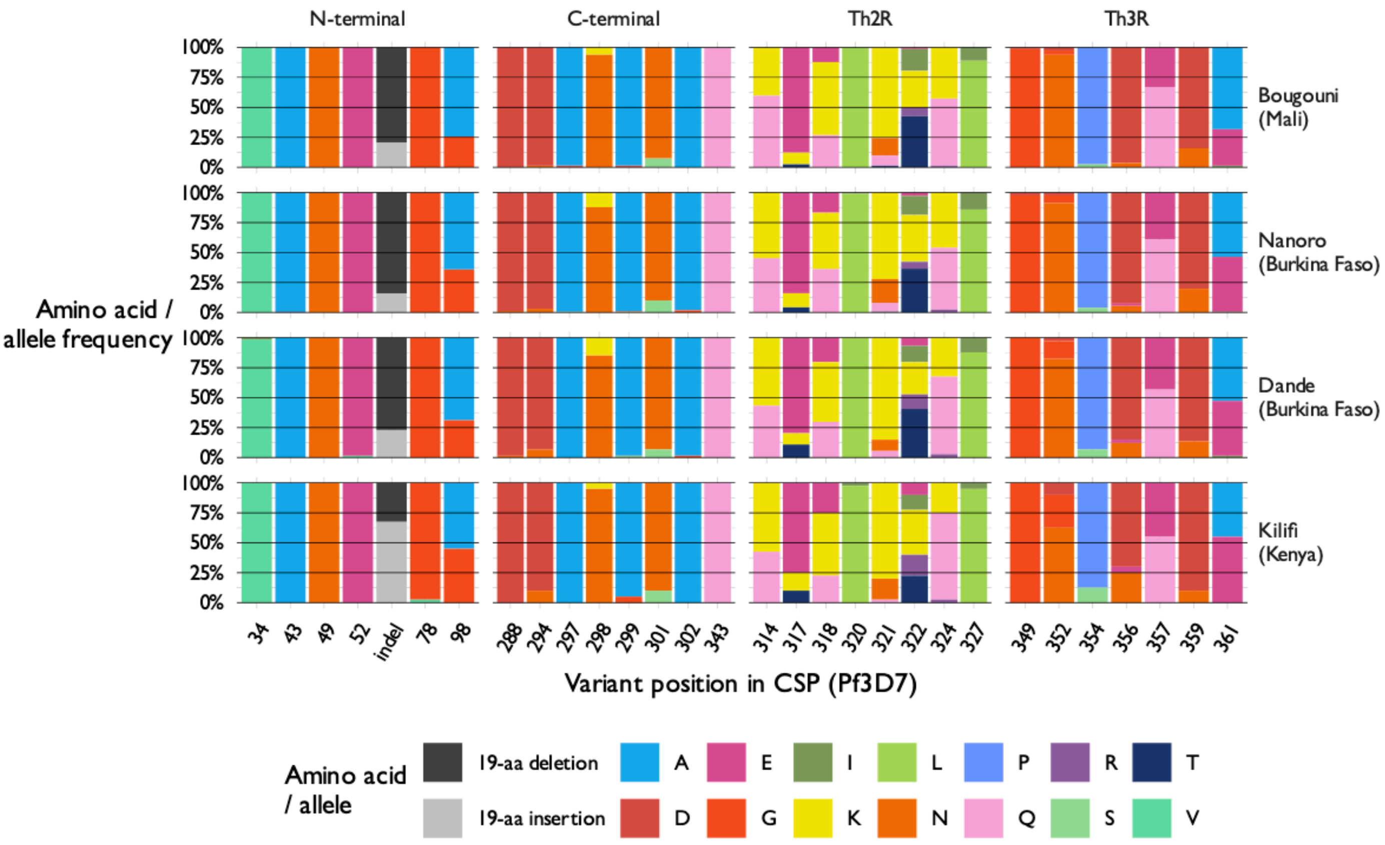

**Figure S 7 Amino acid alleles frequencies of 30 polymorphic residues observed in CSP.**

Plot shows the alleles frequencies (bar height) for each amino acid or allele (colours) observed at each position in the CSP multiple sequence alignment (Figure 5) where cross-haplotype variability was observed. Variant sites are separated by their position in the gene (N-terminal, Th2R, Th3R or other C-terminal region; columns) and frequencies are shown by study site (rows). Variant coordinates are shown at the bottom of the plot and specified with respect to the Pf3D7 gene sequence, also shown in **Figure 5**.

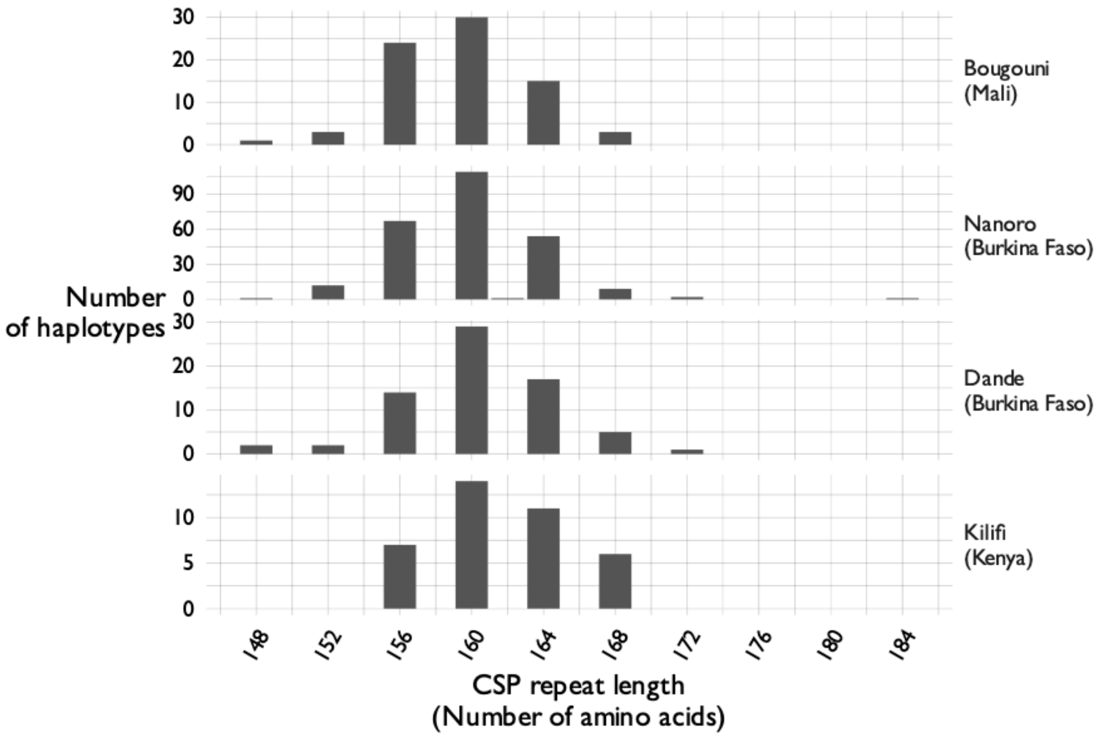

**Figure S 8 CSP repeat lengths across four study sites.**

Plot shows the counts of inferred haplotypes (y axis) having the given CSP repeat length (x axis, measured in amino acids). All but one inferred haplotype has repeat length a multiple of 4; the exception is a haplotype of length 162 observed in Nanoro & Sigle. Four haplotypes (for 2Ry-3dZ-tEm, T89-QKP-wOU, yJW-WTR-ETs and 3HU−5b8−I0d) had no repeat segment or especially short repeats and are excluded from this plot; see main text for a description of these samples.

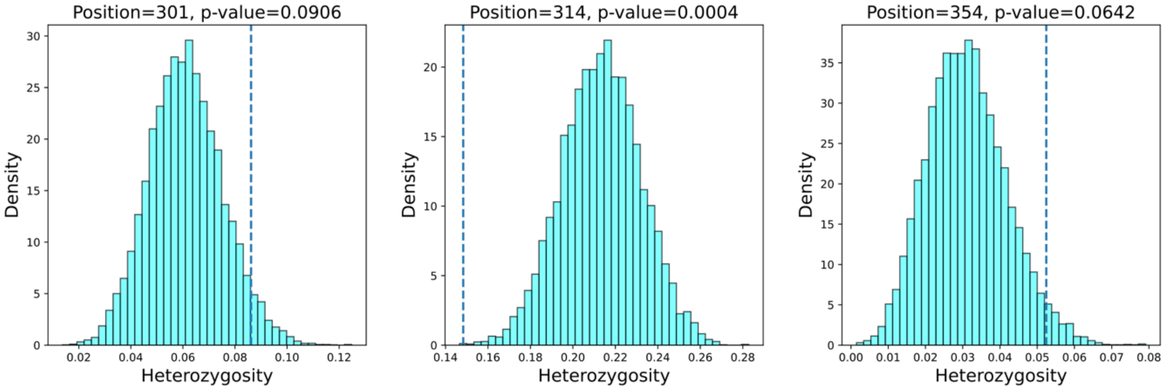

**Figure S 9 Observed and simulated within-infection heterozygosity at three CSP amino acid positions.**

Panels show the distribution of within-infection heterozygosity at *CSP* C-terminal amino positions 301, 314, and 354 computed from 10,000 simulations (rectangles), and the observed within-infection heterozygosity (vertical dashed line), in the N=97 samples with at least two inferred haplotypes at *CSP*. Simulations were peformed by repeatedly sampling haplotypes to make up polyclonal infections matching the observed distribution of haplotype counts and frequencies, with replacement from a total set of haplotypes weighted by frequency observed at the locus. An empirical 2-tailed P-value computed from the simulations is stated above the plot.

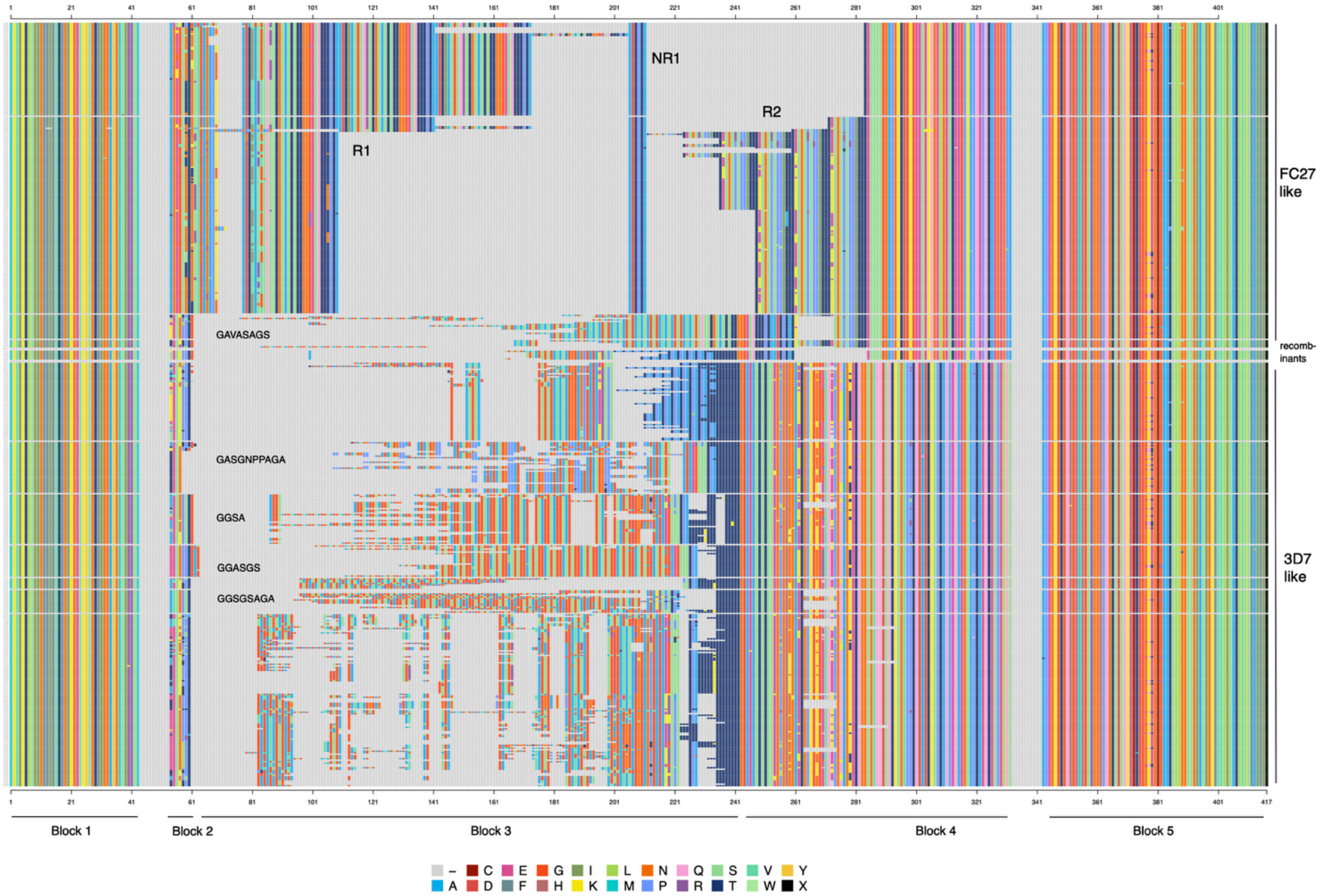

### Figure S 10 The exceptional diversity of *PfMSP2*

Plot shows a manually edited multiple sequence alignment of inferred amino acid haplotypes at the *MSP2* locus. Sequence name annotations (right), FC27-like repeat names, and block identifiers (bottom) correspond to those reported in Ferreira et al ( 10.1016/j.exppara.2006.05.003). We have also annotated consensus sequence for 3D7-like repeat units, where this is clearly identifiable in the alignment.
